# REalist Synthesis Of non-pharmacologicaL interVEntions for antipsychotic-induced weight gain (RESOLVE) in people living with severe mental illness

**DOI:** 10.1101/2024.07.25.24310979

**Authors:** Maura MacPhee, Jo Howe, Hafsah Habib, Emilia Piwowarczyk, Geoff Wong, Amy Ahern, Gurkiran Birdi, Suzanne Higgs, Sheri Oduola, Alex Kenny, Annabel Walsh, Rachel Upthegrove, Katherine Allen, Max Carlish, Justine Lovell, Ian Maidment

## Abstract

**Introduction:** Antipsychotic medications are used to treat individuals with severe mental illness (SMI), but are associated with rapid weight gain and several physical and mental risk factors. Early, proactive weight management is necessary to pre-empt these risk factors. The aim was to understand and explain how, why, for whom, and in what contexts non-pharmacological interventions can help to manage antipsychotic-induced weight gain.

**Methods:** Realist review to identify contextual factors and underlying mechanisms associated with effective, non-pharmacological weight management interventions for adults > 18 years old. Practitioners and lived experience stakeholders were integral.

**Results:** 74 documents used to construct programme theory and 12 testable context-mechanism-outcome configurations. People with SMI benefit from support when navigating interventions aimed at managing the weight gain. From a practitioner perspective, a good therapeutic relationship is important in helping people with SMI navigate early diagnosis and treatment options and facilitating exploring any pre-existing issues. Interventions that are flexible and tailored to the needs of individuals, ideally starting early in a person’s recovery journey are likely to yield better results. Additional sources of support include family, friends and peers with lived experience who can help individuals transition to autonomous goal-setting. The review findings also emphasise the significant effect of stigma/dual stigma on individuals with SMI and weight gain.

**Conclusions:** Successful interventions are collaborative, flexible and underpinned by early and comprehensive assessment with use of appropriate behaviour change approaches. The therapeutic relationship is key with a de-stigmatising approach required. A realist evaluation with primary data is currently underway.

**Practitioner Points:**

- Individuals with severe mental illness on antipsychotic medications are at high risk for rapid weight gain and associated adverse mental and physical outcomes.
- Early comprehensive assessment by knowledgeable, respectful practitioners promotes therapeutic relationship development and identification of individuals’ specific risk factors and support, such as pre-existing disordered eating behaviours and the presence of family/carer and peer support.
- Case management or care coordination needs to be strengthened to ensure individuals’ access to consistent primary and secondary services, as well as community-based services.

## Introduction

Antipsychotic medications are typically used in the treatment of severe (also called serious) mental illness (SMI) (e.g. psychotic disorders, bipolar disorder and major depressive disorder with psychotic experience). Newer, second-generation antipsychotic medications, such as olanzapine, have fewer extrapyramidal side effects, but they are associated with rapid and potentially significant weight gain.^1^

People with SMI die prematurely; the life expectancy, of people with psychosis is reduced by 20 years.^2^ A significant factor in the reduced life expectancy is the complications of comorbid diabetes and heart disease.^2^ Anti-psychotics are associated with weight gain and cardiometabolic risk factors that contribute to the risk of long term physical health conditions including obesity, and cardiovascular and cardiometabolic diseases.^2^

Antipsychotic medication initiation for first-episode psychosis can lead to a 7% increase in body weight within the first year of treatment.^3^ Rapid weight gain in the first 6 weeks is a predictor for long-term weight management challenges due to antipsychotic medications.^4^ Antipsychotic medications promote weight gain primarily by adversely affecting appetite regulation, but also by decreasing physical activity levels (e.g. fatigue) and altering the gut microbiome.^4^

Weight gain can also create a dual stigma for individuals with SMI; the stigma of weight gain and obesity in addition to the stigma of the mental illness.^5^ This may exacerbate some of the underlying psychotic symptoms, such as paranoia, and increasing the risk of suicidal behaviour and other psychiatric disorders, such as anxiety and depression.^6^

Weight management programmes for the general population focus on reducing energy intake via nutrition education, increasing activity levels and teaching behaviour change strategies. These non-pharmacological interventions have demonstrated short-term and long-term effectiveness.^7-9^ Several countries’ clinical practice guidelines for adults with obesity recommend individualised plans for holistic care that address root causes of weight gain and provide support for sustainable behaviour changes.^10-12^ These clinical practice guidelines emphasise preventive strategies and in psychiatric populations, early use of non-pharmacological approaches to manage obesity is recommended.^13^

In the population of individuals with SMI and weight gain from antipsychotic medications, engagement with mainstream weight management programmes is limited due to factors such as low energy, dual stigma and lack of social support.^14,15^ Non-pharmacological interventions to treat antipsychotic medication weight gain for individuals with SMI have been conducted, but they have had mixed results with respect to clinical and cost effectiveness.^16^ Many of these intervention studies were randomised controlled trials (RCTs) of limited duration with specially assigned resources and different outcomes indicators.^17,18^ Given equivocal results from RCT findings, this realist review was conducted to provide a more in-depth look at the contextual factors and underlying mechanisms associated with effective non-pharmacological interventions for weight gain in individuals with SMI on antipsychotic medications.

A realist approach was used to answer the following questions: what types of non-pharmacological interventions are most effective for addressing weight gain from antipsychotic medications in individuals with SMI? What intervention characteristics are associated with intended outcomes? What underlying mechanisms are necessary to create the causal links between specific intervention characteristics and intended outcomes?

Realist approaches are used to construct a testable (i.e. it may be confirmed, refuted or refined) programme theory of what works under what circumstances for whom, as well as why specific interventions or services are effective or not. ^19^ The overall aim of RESOLVE was to understand and explain how, why, for whom, and in what contexts non-pharmacological interventions can help to manage antipsychotic-induced weight gain.

## Materials and Methods

The methodology of realist research was developed to address the question ‘what works, for whom, in what circumstances, and how?’ in different interventions.^19^ RESOLVE is a realist study with two phases: a review based on secondary data (the focus of this paper), and an evaluation phase based on primary data (in progress).^20^ Our research team, with input from our stakeholders, decided to focus on individuals with SMI who live in community settings (e.g., independent living, with families/carers, in supportive housing) where the vast majority of this population live. For community-dwelling individuals with SMI, healthcare services are typically obtained through primary care services (e.g., general practitioners [GPs]) who do referrals to secondary services. Other access may be via community mental health teams. This review encompasses an over-arching programme theory of effective non-pharmacological interventions for individuals with SMI and weight gain from anti-psychotic medications; and context-mechanism-outcome configurations (CMOCs) that provide more granular causal explanations of “how” to plan care and how to implement interventions in community settings for this population.

The methods for the whole project are more fully described in the published protocol^20^, but, for the realist review, the authors used a five-step process:

### 1. Developing the initial programme theory

We used several processes to iteratively develop our initial programme theory:

- Initial scoping searches identified existing eighteen published systematic reviews of non-pharmacological interventions,^14,21-37^ which were used to determine commonly identified factors that were identified as affecting intervention outcomes
- We formed stakeholder advisory groups: a practitioner group and a lived experience group comprised of individuals with SMI and weight gain from antipsychotic medications and their informal family carers. The McPin Foundation, UK, was a partner on our research grant, organizing the lived experience group members for this project. The practitioner group was organized by the lead researcher (JH) with assistance from research team members. Both stakeholder groups provided insights into important concepts to explore in the literature and met up regularly throughout the course of the project.
- We drew on the collective experience within the project team.

Using the processes outlined above, we iteratively constructed an initial programme theory of what a successful recovery journey might look like for an individual with SMI and weight gain from antipsychotic medications.

### 2. Searching for evidence

To avoid duplicating the extensive work of previous review teams, we gathered documents by consulting the lists of included studies for the three most recently published reviews that focused on weight management within SMI^14,30,36^ (n=80 studies from the papers). We subsequently used CLUSTER searching techniques^38^ to identify companion or ‘sibling’ papers, related to the studies identified for inclusion. We followed up reference lists of included studies and searched for other documents associated with these studies using the study names, ID numbers (e.g. clinical trial registration numbers) and by searching for author names. We sought to identify related qualitative studies, process evaluations, protocols and other documents likely to contain further details on important contexts and potential mechanisms at work in the interventions under study.

Our formal search strategy was developed by an academic information synthesis specialist, in consultation the wider project team and combined free text and subject heading terms for SMI, terms for non-pharmacological interventions and terms for weight-related outcomes of interest. It was translated for use across eight bibliographic databases (MEDLINE, Embase, PsycINFO, CINAHL, Cochrane Library, Scopus, Web of Science Core Citation Indexes (SCIE, SSCI, SHCI, ESCI, CPCI, BKCI) and Sociological Abstracts) with the aim of achieving broad disciplinary coverage. Our protocol indicated that we would also run searches in Google Scholar and search for grey literature, but this was deemed unnecessary following screening, in light of the volume of relevant literature already retrieved.

The main searches were conducted in February 2022. Additional articles were also identified via Google Scholar alerts from February 2022 through to August 2023. Searching was conducted in an iterative fashion, depending on ongoing feedback from the stakeholder groups. For example, the issue of food craving and binging was raised by individuals with lived experience, and documents were searched and included in the review to address pre-existing disordered eating behaviours. Searching continued until sufficient data were located to refine the initial programme theory and CMOCs. Supplementary File 1 contains the full search strategy.

### 3. Selection, appraisal and data extraction

Documents were selected for inclusion in the review in multiple steps.

First, screening of titles and abstracts was performed in RAYYAN, a web-based platform which supports reviews. A 10% sample of documents was screened in triplicate by three members of the project team. The inclusion and exclusion criteria are outlined in Table 1.

**Table 1.**
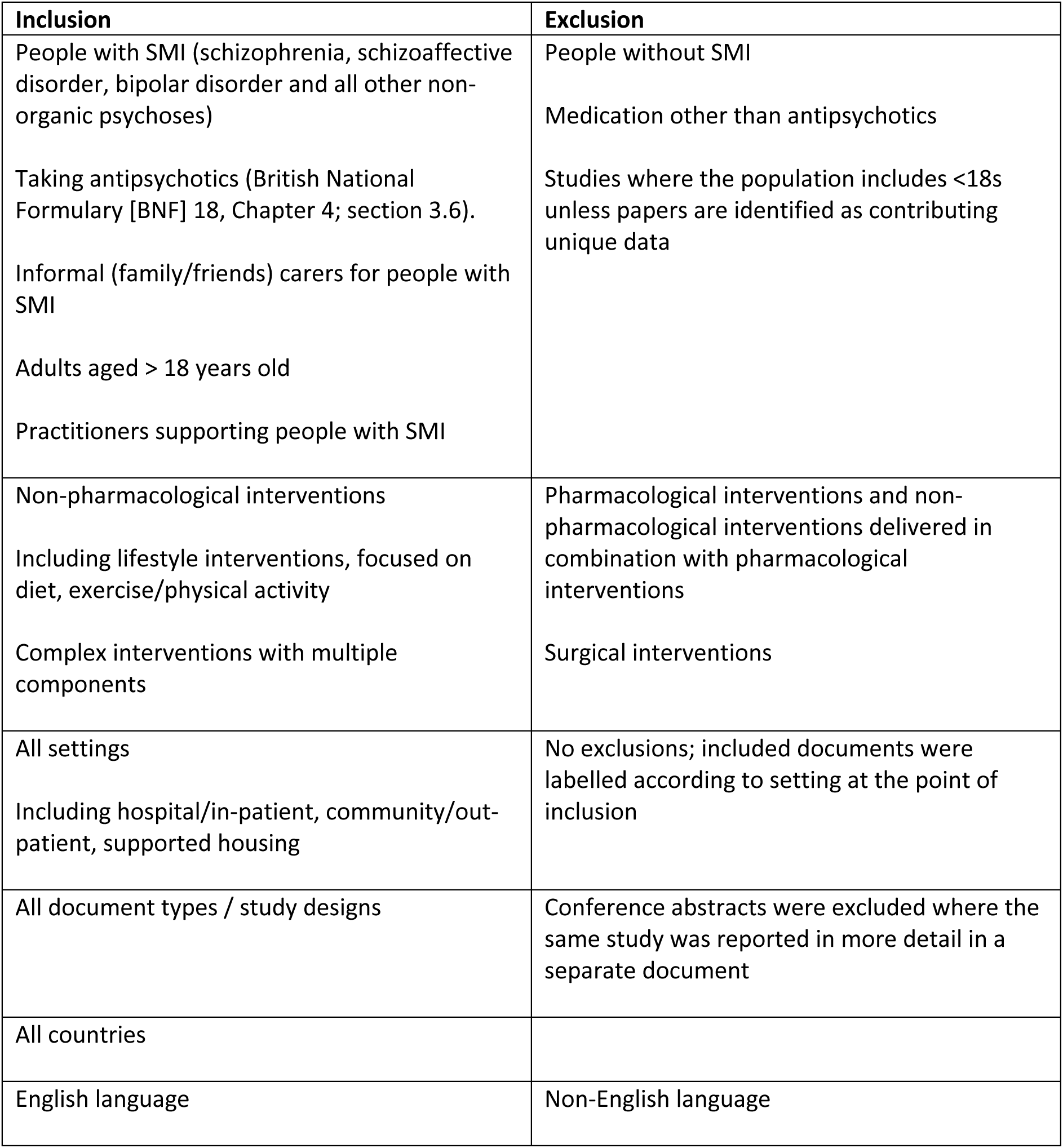
Inclusion criteria and exclusion criteria used for abstract and full text screening.

Second, a full text screen of remaining documents against the same inclusion and exclusion criteria was conducted. A ‘star rating’ system which ranked and prioritized documents according to relevance was adopted.^39^ At this stage the decision was made to focus on the community setting due to the volume of data retrieved, differences in settings between inpatient and community, and because most people with SMI are in the community. Following a pause in the review due to staffing changes, the ‘five-star’ and ‘four-star’ papers were rescreened to determine potential contribution to the refinement of the initial programme theory. Final documents were selected and coded based on their relevance, richness and rigour. Relevance is based on document content. Are there sections of the document that contain data that are relevant to theory and CMOC development? Richness is defined as whether the document contains content pertinent to Cs (Contexts), Ms (Mechanisms) and Os (Outcomes), and whether the documents have clear C-M-O linkages. Rigour is based on the trustworthiness of the data.^19^ Figure 1 is the PRISMA diagram of the inclusion/screening process. Supplementary files 2 contains the characteristics of all 74 documents included in the review.

**Figure 1.**
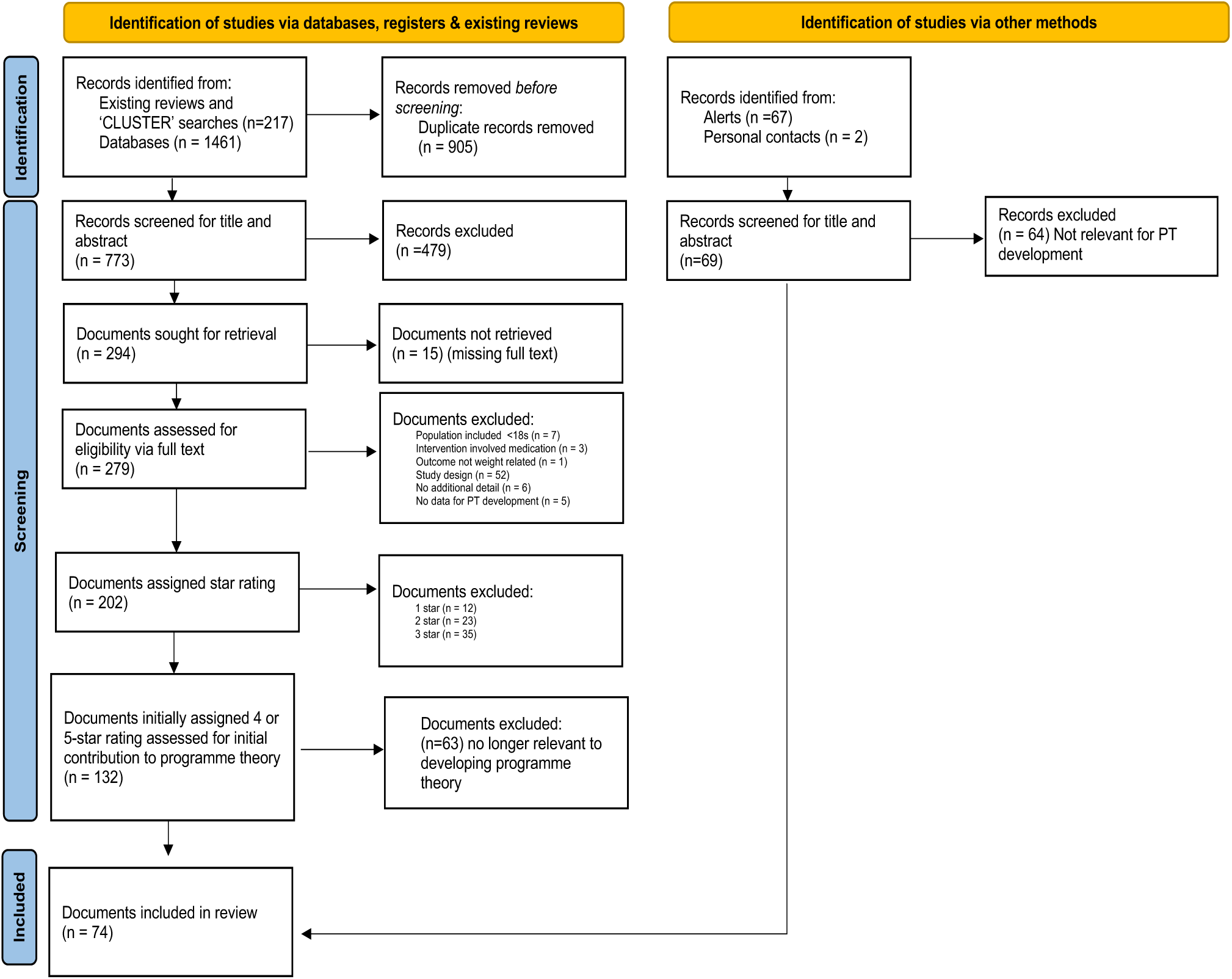
PRISMA flow diagram.

### 4. Data Extraction and analysis

Key characteristics of documents were extracted and imported into Microsoft Excel. Included documents were uploaded into NVivo and relevant extracts of data were extracted and coded. Coding was both inductive and deductive. A coding framework outlining conceptual categories was established by reviewers and documents were coded according to their identification of key contextual factors associated with intended outcomes (i.e. effective weight management). In addition, documents were coded for potential underlying mechanisms related to ‘why’ specific contextual factors were necessary to produce intended outcomes.

### 5. Data synthesis, CMOC development and programme theory refinement

Coded data were used to develop CMOCs and to refine the initial programme theory. The realist researchers met regularly to extract potential Contexts-Mechanisms-Outcomes from each conceptual category. Initial CMOCs with corresponding quotes from review documents were shared monthly with the entire research team. At these meetings, the CMOCs and the programme theory were refined and validated by our team’s subject matter experts and lived experience group members. A significant contribution of subject matter experts and the lived experience project team members was confirmation of resonance with their own lived experience and replacement of any blaming or shaming language with neutral, non-stigmatizing language.

This review followed the RAMESES realist review quality and publication standards.^19^ An important feature of realist approaches is the inclusion of stakeholder groups to provide feedback and advice throughout the review process.^40,41^ Our stakeholder groups and the research team made a decision to develop the theory and CMOCs from the perspective of individuals with SMI and medication-induced weight gain (versus practitioners), since ultimately, effective weight management depends on how interventions and support work for individuals in the real world. The RAMESES checklist for realist reviews can be found in Supplementary File 3.

## Results

The initial search strategy which included the documents identified from three recent reviews, cluster searches and academic databases identified 773 unique results. 279 documents were read in full-text, with 132 documents being assigned a four or five-star rating. Following a re-appraisal of documents, in light of the developing programme theory, 63 documents were excluded, leaving 69 four and five-star papers included in the review. An additional 5 papers were identified via google alerts and personal contacts giving rise to 74 papers in total. Our searching and screening processes are detailed within the PRISMA diagram in Figure 1.

Of the papers reporting primary data, twelve were RCTs^42-53^, with four additional studies reporting secondary analyses from existing RCTs.^54-57^ One RCT reported a follow-up after a two-year period.^58^ Five papers were pilot studies,^59-63^ and four were feasibility studies.^49,64-66^ Other study designs included cross-sectional studies,^67,68^ experimental action research,^69^ non-randomized clinical trial,^70^ and observational ^71^ or quasi-experimental observational.^72^ Twenty-six qualitative papers were included, with 24 presenting intervention study results,^51,73-95^ the two remaining qualitative papers reported mixed methods (including qualitative) evaluation of the implementation of a RCT,^96^ and a reflection on the delivery of a RCT.^97^

The remaining papers included: three protocols,^98-100^ four papers that contained methodological descriptions of intervention development,^101-104^ five literature reviews,^105-109^ two commentaries^15,110^ and a report.^111^

Studies were conducted in different countries. Six papers were from Australia,^81,84,88,104,106,109^ two from Belgium,^112,113^ three from Canada,^68,92,107^ one from Denmark,^58^ and one was based more broadly in Scandinavia.^71^ One study was conducted in Lebanon,^67^ and another in the Netherlands.^103^ Four papers were from Sweden.^48,69,75,76^ Thirteen papers were from the United Kingdom.^15,70,80,82,83,87,96,98,99,101,102,114,115^ The remaining forty-two papers were from the United States of America.^42-47,49,50,52-57,59-63,65,66,72-74,77-79,85,86,89-91,93,95,97,100,105,110,111,116,117^

### A Program Theory of Non-pharmacological Interventions for Individuals with SMI and Weight Gain from Antipsychotic Medications

Given the mixed results from many of the intervention studies, particularly the RCTs, programme theory construction and CMOC development were primarily based on documents with participant and practitioner descriptions of effective non-pharmacological interventions for antipsychotic-induced weight gain. Through ongoing stakeholder group discussions and subject matter experts’ input, we refined the following <u>programme theory</u>:

The recovery journey for individuals with SMI and medication-induced weight gain begins with the development of trusting, therapeutic relationships with practitioners. The therapeutic relationship is where holistic assessment of mental and physical needs and co-developed goals can be pursued by individuals with their practitioners. Individuals need to be informed and aware of the deleterious side effects of anti-psychotic medications, particularly the impact that rapid weight gain can have on their health and wellbeing. A de-stigmatising approach is needed throughout individuals’ recovery journey to overcome the dual stigma of weight gain and SMI that adversely affects individuals’ physical and mental health, their self-efficacy and willingness to participate in weight management programmes. Effective weight management programmes provide flexible offerings of nutrition planning, physical activities and behaviour therapies via individual or group sessions. Phased approaches, include an initial intensive phase to better prepare individuals for programme engagement, followed by a longer, second maintenance phase where individuals learn how to monitor their own health data and to set small, manageable behaviour change goals with support from skilled practitioners, trained facilitators and/or peer mentors. Easy-to-use tracking and self-monitoring devices are a useful source of objective data to raise individuals’ awareness of behaviour change progress. During the third, transitional phase, individuals take control of their own health behaviours, which includes weight management. Given the number of physical and mental barriers associated with SMI, additional support from families/carers, friends and peers contribute to individuals’ sense of achievement and belonging. Learning from peers with lived experience is especially important for raising hope for a positive future.

### The Context-Mechanism-Outcome Configurations with Accompanying Citations and Extracted Quotes

Table 2 contains the 12 CMOCs (Context-Mechanism-Outcome-Configuration) that underpin the programme theory. The table includes references for all the papers included in this review which underpinned each CMOC. A table of CMOCs and illustrative evidential quotes can be found in supplementary file 4. The CMOCs are organised for practitioner use, beginning with therapeutic relationship development between individuals and practitioners, to characteristics of effective interventions and support.

**Table 2.**
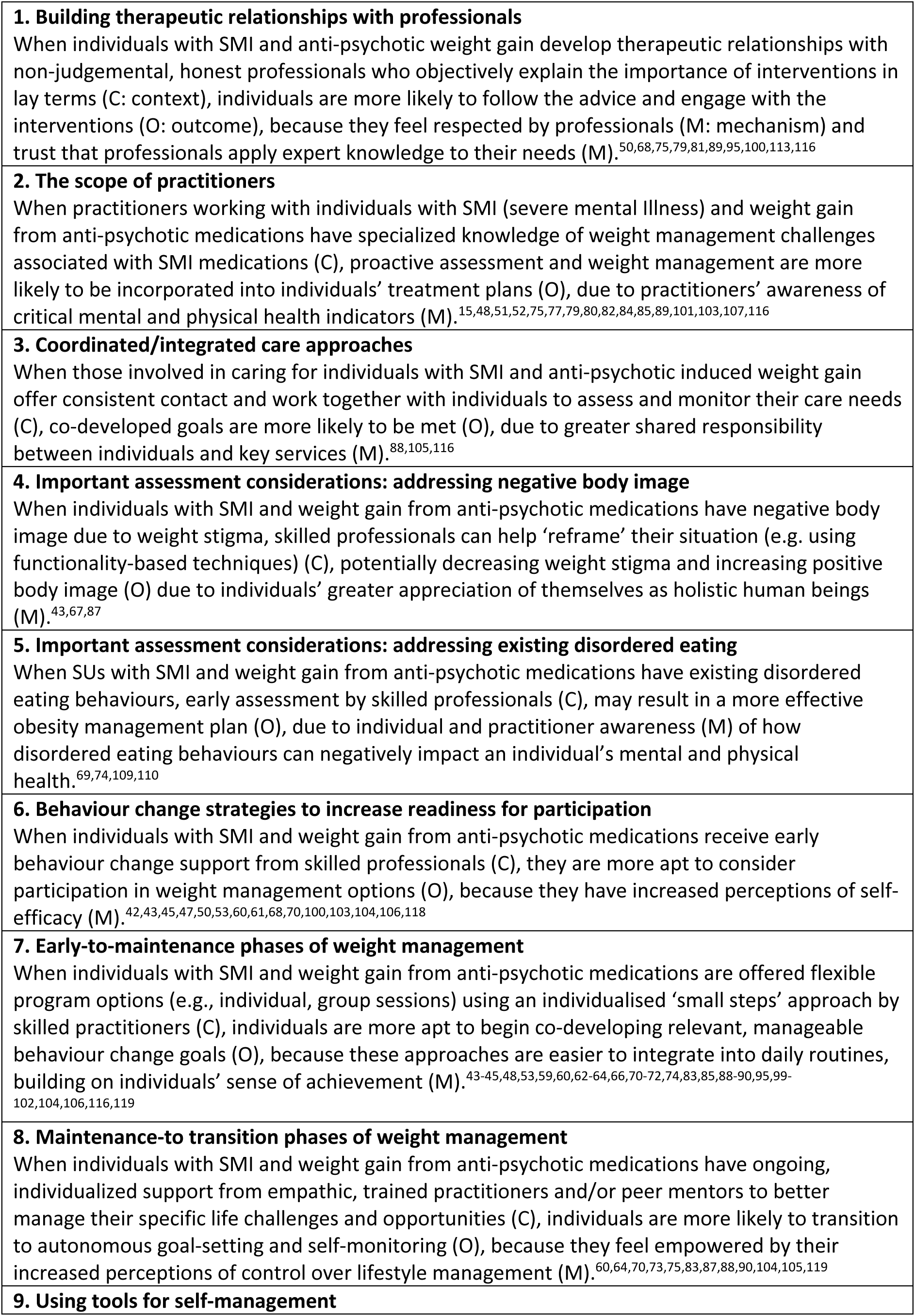
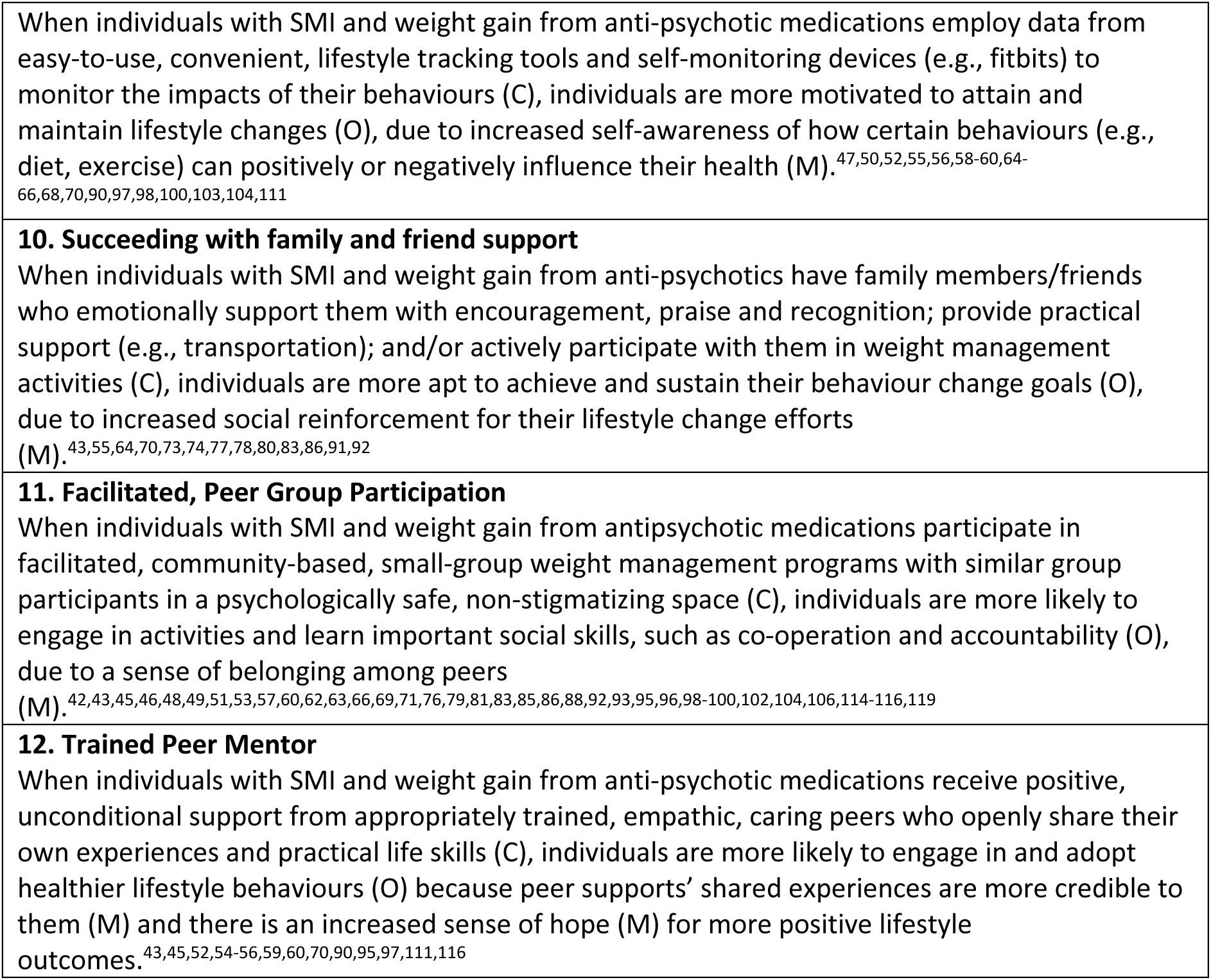
Context-Mechanisms-Outcomes configurations.

The first five CMOCs pertain to the initiation of a therapeutic relationship between community-based practitioners and individuals with SMI and medication-induced weight gain. The therapeutic relationship is an important social context for individuals with SMI, where they learn about their illness, prescribed antipsychotic medications, medication side effects and realistic management of their health needs.

In CMOC 1, the contextual factors of importance to individuals are practitioners who are non-judgmental and honest with them about the side effects of their antipsychotic medications. Because weight gain can be rapid, information needs to be conveyed early on in repeated, concrete, simple language that individuals can understand and relate to their own life circumstances.^116^ The underlying mechanisms associated with relationship formation are individuals’ perceptions that their opinions are respected (mechanism) by practitioners and trust (mechanism) in practitioners’ advice to them.

Because the therapeutic relationship is a gateway to individuals’ engagement in co-development of a weight management plan, a significant problem in primary care services is the scope and time/availability of practitioners. As detailed in CMOC 2, specialised knowledge and relational skills are needed by practitioners working with individuals with SMI and weight gain in community settings. Practitioners need to be aware (mechanism) of holistic screening and assessment for this specific population—to ensure more proactive management of weight gain for individuals with SMI.^15^

A number of different, community-based care models have been piloted to provide more holistic assessment and management of individuals’ physical and mental health needs. As described in CMOC 3, key contextual characteristics of holistic care are collaboration and coordination across services. One mental health collaborative care model (CCM) included in our review documents was based on Wagner’s chronic care model.^105^ The CCM emphasises connections between mental health specialists and general practitioners, facilitated by a case/care manager, such as a mental health nurse. Because continuity and trust relationships are important to individuals with SMI and weight gain, the case manager is often the consistent contact between the team and individuals. Another key characteristic of the CCM is measurement-based care: mental–physical health assessment tools are systematically used to track weight management progress, and the case manager is often responsible for monitoring, collecting, and sharing data with the team and individuals. Shared data are an important means of ensuring individuals’ understanding of how their individualised weight management plan is making a difference to their quality of life.^105^

Shared responsibility (mechanism) ensures effective, collaborative care across services, including engagement and collaboration with individuals. When the research team discussed this CMOC with the lived experience group members, the group members highlighted how siloed care often arises when no one takes responsibility for their role in complex care situations. The lived experience group members felt that services/practitioners need to accept their shared responsibility in care coordination and delivery. The lived experience group also stated that true person-centered care requires active engagement with individuals–beginning with assessment and continuing on to individuals’ independent control of their care decisions.

The following CMOCs – 4 (negative body image) and 5 (disordered eating behaviours) – acknowledge specific assessment components for individuals with SMI and weight gain. Functionality appreciation techniques are designed to raise individuals’ appreciation (mechanism) of what their bodies are capable of doing. Focusing on body functionality (i.e. the body positivity movement) versus appearance is one way to positively ‘reframe’ how individuals view themselves.^67,87^ Another assessment area is the presence of disordered eating behaviours, which are more common in people with psychosis than the general population.^109^ A systematic review of disordered eating behaviours in individuals with schizophrenia spectrum disorder reported rates of 4.4% to 45% for binge eating, 16.1% to 64% for food craving, 27% to 60.6% for food addiction, and 4% to 30% for night eating.^109^ Therefore, early assessment and management of pre-existing disordered eating behaviours can heighten individual and practitioner awareness (mechanism) of intervention options for holistic weight management.^110^

Barriers to participation in weight management interventions include the dual stigma of SMI and weight gain and the cognitive deficits and fatigue associated with SMI.^43,100^ As stated in CMOC 6, brief behaviour change interventions, such as motivational interviewing (MI), can be used to raise individuals’ self-efficacy beliefs (i.e. ‘I can do it’) and readiness to participate in weight management planning and treatment. ^43^ Interventions often fail when individuals are not open to participation.^75^ Motivational interviewing is one ‘guiding style’ and a relational, strengths-based approach that foregrounds individuals’ existing strengths and future aspirations. Furthermore, MI is a technique that practitioners can readily learn and apply.^100^ Therefore, prior to the start of any intervention behaviour change strategies, such as MI, should be used to raise individuals’ sense of self-efficacy (mechanism) and readiness to participate.^87^

CMOCs 7 to 9 cover key characteristics of effective weight management interventions for individuals with SMI. In CMOC 7, during the early intervention phase, a primary focus is individual-practitioner co-development of small, manageable behaviour change goals that can be integrated into individuals’ daily routines and social and home settings. Successful interventions are based on a few, specific skills which are broken into smaller units and repeated as needed. Learning aids, such as visual cues,^100^ are also used to reduce memory and attention load and to improve adaptive functioning. Small, progressive successes are more apt to build SUs’ sense of achievement (mechanism),^64,75,95^ and there is some evidence that a ‘small steps’ approach is more effective in sustained long-term behaviour change.^95^

CMOC 8 is a continuation of ongoing, collaborative goal-setting between individuals and trained practitioners and/or peer mentors. During the maintenance phase, individuals learn how to monitor their own progress and set their own behaviour change goals based on continued, individualised assistance with identification of potential/actual life opportunities and challenges. The transition phase is marked by individuals’ capacity to take control of their own health behaviours as a result of their feelings of empowerment (mechanism). Empowerment represents how individuals feel when they have mastered the ability to enact valued health behaviours.^70^

As stated in CMOC 9, easy-to-use tracking and self-monitoring tools may give more control to individuals as they learn to establish their own, small goals based on their increased self-awareness (mechanism) of how their health behaviours are associated with positive or negative health outcomes. As noted in one qualitative study of individuals with SMI and weight gain, when individuals began monitoring their caloric intake, they were surprised by what they learnt about their food choices. Raised awareness helped them develop healthier consumption habits.^97^

CMOCs 10 to 12 describe the key types of support described in the review literature. With psychosis, early and intensive social support are often needed to help individuals develop a greater sense of self-efficacy and readiness to change. Ongoing reinforcement is important to sustain positive behaviour change. A primary source of emotional support is the family,^64,73,75^ although not all family members may be accessible or helpful. In an initial assessment, practitioners need to determine the individual’s sources of support, particularly the role of family and peers.^64^ In one intervention for obesity management, the US Fit Together programme, education and activities were designed for individuals and their family members or friends. The individual participants stated that diet and physical activity were easier to carry out with a family member who was also committed to improved health goals.^64^ The outcomes were more positive and sustainable for individuals when their family members also succeeded in losing weight and making behaviour changes.^64^

Individuals with SMI often have difficulty in establishing and maintaining social relationships.^73^ Because family members (where available) are a key source of support for individuals (e.g. emotional, social, and material),^64^ practitioners should also determine – working with family and significant others – what types of resources and social reinforcement (mechanism) family/friend support need to provide optimal support, such as signposting to services or social prescribing contacts.^80^ Some review papers recommended that families and significant others should receive information and skills training on coping with SMI, as well as alerting them to possible medication side-effects.^77,78^

Individuals learn from and adopt others’ behaviours through social interactions, where observation, experience and positive reinforcement help to shape individuals’ decisions and actions. Many group interventions, such as group-based weight management sessions, are built on the powerful influence of peer social support and a sense of belonging (mechanism).^55^ As noted in CMOC 11, facilitated sessions with similar group participants creates a non-stigmatising, psychologically safe space where individuals can openly share their particular needs and challenges with each other.^79,96^ In group-based interventions with high levels of peer interaction, participants reported a greater sense of co-operation while learning together,^88^ increased knowledge of social roles,^62^ and a greater sense of social accountability.^81,93^

The final CMOC 12 represents a trend of using trained peer mentors with lived experience (i.e., of SMI and weight gain from antipsychotic medications) to facilitate group sessions, and to be available as needed for one-on-one support. Peer mentors may be another cost-effective strategy for promoting positive behaviour change in this population.^59^ The capacity of peer mentors to truly empathise with participants’ recovery journeys was emphasised in the review documents. In the US PeerFit program, trained peer mentors were able to explore participants’ past and present ‘readiness to change’ behaviours, based on their capacity to relate to programme participants’ life stories.^59^ In this intervention, a collaborative coaching and learning model was used, where a trained facilitator offered one-on-one counseling, while the peer mentor role modelled healthy behaviours and provided assistance and support with individuals’ goal-setting. Peer mentors who were interviewed at the completion of the study stated that they preferred reciprocal, non-hierarchical relationships with participants; the coaching role made them uncomfortable.^59^ Another intervention using trained facilitators and peer mentors reported how both types of support complemented each other.^116^ In a related paper, interviews were conducted with programme participants who clearly differentiated between the characteristics of practitioners in facilitator roles (referred to as non-peer facilitators) and trained peer mentors (referred to as peer support).^90^ Practitioners were described as task-oriented and goal-focused, while peer mentors were more positive and relational—thus building a sense of hope (mechanism) and the possibility of positive change. Given their own lived experiences, peer mentors were better able to contextualise how to make small and manageable lifestyle changes. Peer mentors were also described as providing unconditional and unwavering support. In this intervention, supervisors of peer mentors valued peer mentors’ ability to ‘normalise slips’, such as a food binge or weight gain, and to provide credible examples of their own, very human struggles with non-healthy choices.^90^

## Discussion

### Key findings

Antipsychotic related weight gain, as highlighted in the introduction, poses a serious risk to the physical health of those with SMI and it should be a top priority for healthcare related policy. Indeed, callers to a mental health charity helpline reported weight gain resulting from antipsychotic medication as the most distressing side effect.^120^

This realist review is comprised of a programme theory of weight management for individuals with SMI and weight gain from antipsychotic medications, underpinned by 12 CMOCs with applicability to practitioners caring for this population in community settings. CMOCs 1-8 focus on individual-practitioner therapeutic relationships. Initial relationships are based on practitioners’ nonjudgemental attitudes, honest, objective information-sharing, and individuals’ trust in practitioners’ professional advice (CMOC 1). Early, comprehensive assessments, with an understanding of the challenges of managing anti-psychotic weight gain, need to include the exploration of pre-existing issues, such as negative body image and disordered eating behaviours (CMOCs 2,4,5). Given the complexity of individuals’ physical and mental health needs, coordinated and integrated care approaches are effective means of care delivery for this population (CMOC 3). CMOCs 6-7 highlight the need for flexible programme options and individualized “small step” approaches, ideally starting from an early point in recovery, for easier integration of behaviour change goals into individuals’ daily routines. CMOC 8 acknowledges how practitioners and peer mentors can assist individuals with transition to autonomous goal-setting, while CMOCs 9-12 offer details on non-practitioner support, including the use of self-management tools (CMOC 9), family and friend support (CMOC 10), peer group participation (CMOC 11) and trained peer mentor support (CMOC 12) A unique aspect of this review is a theory with testable CMOCs that suggest the intertwined nature of the SMI recovery journey and effective weight management.

### Comparable Literature to Support the Programme Theory and CMOCs

This section compares our findings to the wider literature, including the non-SMI literature. However, compared to the non-SMI population there are likely to be factors that make managing weight even more challenging in people with SMI including fears about medication non-adherence, mental health stigma, the negative symptoms of schizophrenia, the lack of trust between clinicians and service users and the roles and responsibilities of primary versus secondary care.^121-123^

The importance of supportive non-judgmental therapeutic relationships between service users and the healthcare professionals who are responsible for their care is vital; previous research from the present research team also found that an honest and trusting therapeutic alliance is likely to result in a greater engagement with any interventions suggested.^39^ Due to the rapid onset of weight gain associated with some antipsychotic medication, service users need to be supported at an early stage in their recovery journey to deal with potential consequences associated with their medication regime.^121,124^

Guidelines in this area do exist.^2,125^ For example, annual physical health monitoring for individuals with SMI is recommended by the National Institute of Clinical Excellence,^126-128^ though completion rates are suboptimal and stakeholder discussions report lack of follow-up.^2^ However, previous research has identified the lack of a rigorously developed guidelines focused specifically on the management of antipsychotic weight gain.^129,130^ This can make it challenging for healthcare professionals to offer appropriate evidence-based support to individuals with SMI. Additionally, there is a lack of consensus as to who should be responsible for the physical healthcare of individuals with SMI.^131,132^

Our findings highlight the importance of stigma in managing weight gain for people with SMI who can experience many types of stigma, including societal biases towards individuals who are overweight/obese.^133^ Across the healthcare spectrum practitioners often believe that obesity is due to lifestyle choices and the responsibility for managing the condition resides with individuals.^134-136^ Stigmatising beliefs held by healthcare professionals can negatively influence both the identification of weight gain and obesity as well as onward referrals for managing the conditions.^137-139^ Weight stigma can adversely influence the quality of healthcare delivery and within psychiatry, stigmatising staff beliefs have been identified as a barrier to receiving physical health care.^136,140,141^

In the non-SMI literature, in a UK survey, practitioners cited individuals’ lack of interest (72%) and lack of motivation (61%) as major reasons for not discussing weight with individuals.^142^ Yet 65% of individuals living with obesity wanted their practitioners to discuss weight and related co-morbidities.^142^ Again, in the non-SMI literature, a multi-country survey of individuals with weight gain issues reported healthcare avoidance due to inattentive listening and less respect from practitioners.^143^ Furthermore, weight stigma has a significant, adverse impact on individuals’ social identity, resulting in avoidance of potentially stigmatizing public settings (e.g. gym) and stigma “escape” by engaging in unhealthy behaviours, such as stress-induced eating.^139^ As already noted, there are likely to be additional challenges faced by people with SMI because of the double burden of mental health stigma.

Our findings also highlight the role of supportive practitioner-SU relationships in identifying pre-existing risk factors that can potentiate health risks from antipsychotic medications. For example, existing disordered eating behaviours and negative body image are important components of early assessment and individual-practitioner goal-setting.^144^ The prevalence of pre-existing disordered eating within the SMI population is not well known, however, our lived experience advisory group supported our limited findings and given the additional stigma associated with this practice, stressed the importance of relational practitioner approaches in engaging individuals in honest and difficult conversations^145,146^ so that early identification occurs and appropriate treatment options can be provided. However, the appropriate assessment and treatment of eating disorders in the context of obesity is challenging even in the non-SMI population;^144^ more research is needed to determine best practice.

An individual’s sense of social identity is influenced by their social roles (e.g. partner, brother, son, employee). In socially stigmatizing settings, individuals’ social identity becomes devalued and replaced by a stigmatized view of oneself: Individuals with self-stigma believe they have less self-worth and self-efficacy—resulting in withdrawal and in many instances, self-imposed isolation.^138,147^ Supportive therapeutic relationships with practitioners are considered one, important way to reduce self-stigma.^148^ As identified in our review and discussed in other literature, behaviour change therapies that use relational approaches can build and maintain positive self-confidence and self-efficacy^149^ and positive relationships with families and friends.^147,150^

Given the pervasiveness of weight stigma in society and particularly in psychiatry where individuals may suffer from ‘dual-stigma’ from SMI and weight gain, practitioners are urged to shift in care management from weight-centric to holistic health approaches that recognise that weight loss may not be achievable or sustainable for many individuals.^151^ Practitioners, therefore, need to attenuate stigmatising beliefs and behaviours about weight, and instead promote modifiable health behaviours (versus weight loss) that aim to build more respectful, trusting individual-practitioner relationships.^151,152^

Our review emphasised the importance of supportive approaches that invite honest, transparent conversations to engage individuals and to collaboratively establish realistic whole health goals between practitioners and individuals.^153^ One behaviour change strategy, MI, can be used by practitioners to clarify goals and to establish consonance between goals and current behaviours. This approach is based on person-centred principles that respect individuals’ autonomy and supports their self-efficacy.^154,155^ Acknowledgement of personal goals is more likely to trigger the underlying mechanisms in our review, such as individual’s perceptions of respect from practitioners, trust in their practitioners, a greater sense of shared responsibility, and enhanced self-awareness, appreciation for self as a whole person, self-efficacy, achievement and control over their own health needs.^156^

This review also highlights how social support from others, such as families/carers, peers in group-based interventions and peer mentors have contributed to individuals’ participation and successes in weight management/behaviour change programmes. In the weight loss literature (non-SMI), social influence (e.g., role modeling) and social norms can have a particularly significant impact on eating, for example.^157^ In a US survey study with individuals with SMI, a greater sense of group belonging mitigated against self-stigma or negative beliefs about oneself.^147^ Our review highlights how the presence of good social support and social networks may be critical to the proposed underlying mechanisms of intervention effectiveness including social reinforcement from others, a sense of social belonging, credibility in behavior change offerings and hope in a healthier future.

Our review identified the need for coordinated, collaborative holistic care management to support individuals with SMI in community settings with medication-related weight gain. Currently, a lack of integration across primary care, mental health services, physical health services, which are largely separate, and social care results in fragmented care for people living with SMI, resulting in unmet needs for people living with SMI and weight gain.^123^ Given the potential for complex physical and mental health risks, integrated care may be a preferred delivery model for individuals with complex mental-physical health needs.^123,158^

In the UK, although the prescribing of antipsychotic medication is typically undertaken by psychiatrists residing in secondary care, care of physical health conditions typically resides with GPs in primary care.^159,160^ Those deemed to possess stable mental health may be referred back to the GP for complete management of their physical and mental health.^159,161^ Caring for people with SMI presents unique challenges for primary care practitioners particularly around managing anti-psychotic medication.^161^ With primary care ill-equipped to manage the complexity of SMI and the associated physical health conditions people with SMI may not receive appropriate preventative support from their practitioners particularly with the current pressures on primary care. Likewise, within secondary care psychiatric services, mental health practitioners can often feel ill-equipped to deal with physical health conditions such as weight gain resulting from antipsychotic medications,^162^ meaning that wherever individuals with SMI and weight gain are cared for within the system, they are likely to receive sub optimal preventative care for physical health conditions.

An important healthcare trend noted in our review and the literature is person-centered care. Integrated care teams need to include both practitioners and individuals with SMI and family members.^163^ Physical health monitoring in SMI is likely to need primary care working in tandem with secondary care. ^124,164^ There is a need to develop more effective ways for managing the physical health of people with SMI and integrated care and innovative practitioner collaborations may be one way to address any competency gaps for complex physical-mental health needs. For example, one pilot from Scotland is using clinical pharmacists to assist with physical health monitoring and polypharmacy reviews for SUs with SMI.^165^ Other physical healthcare gaps noted in the literature (which may be remedied with coordinated/integrated care) include regular monitoring for cardiometabolic risk factors associated with antipsychotic medications and weight gain.^13,126,166^ In a systematic review of recommended metabolic risk screening for individuals with SMI, less than 50% of people were being appropriately monitored for one risk factor, and only 11% of this population was meeting recommended monitoring requirements.^167^

### Implications for Practice and Research

Our findings emphasise the importance of providing support for people with SMI and antipsychotic related weight gain. Managers of services should consider how support structures could be incorporated within existing services. Peer support groups provide one such avenue whereby people with SMI could gain positive experiences of managing their conditions through learning from the experience of others in non-judgmental environments.

Individuals with SMI and taking antipsychotic medication need access to practitioners who are knowledgeable about the challenges of antipsychotic related weight gain so that appropriate treatment options may be provided. However, given the siloed nature of healthcare, particularly in the UK, this may be difficult to realise; future research should focus on the organisation of services so that more coordinated and holistic care can be delivered to people with SMI. Our findings also highlight the importance of conducting early comprehensive assessments so that preventative approaches could be adopted. Such assessment should consider the impact of negative body image and disordered eating behaviours on individual’s mental and physical health.

In general, the review found a lack on research on the patient perspective on anti-psychotic weight gain particularly in certain populations such as ethnic minority communities and in learning disabilities. Over-prescribing is also more common in certain ethnic minority groups; as are the metabolic side-effects of anti-psychotics, such as diabetes and total cholesterol.^1,168,169^ People with learning disabilities are 16 times more likely, than the general population, to be prescribed an anti-psychotic.^170^ People with intellectual disabilities have a higher prevalence of obesity and metabolic syndrome.^171,172^ More research is required in these underserved populations.

A major concern of our lived experience group was a lack of literature on the influence of diverse cultures, religious faiths and ethnicities on intervention success. For example, the literature we reviewed did not specifically discuss if/how nutritional interventions were tailored to meet non-Western food preferences. Future research should include a focus on the positive or negative influence of culture on intervention outcomes and how interventions should be adapted to best serve different communities.

### Strengths and Limitations

A key strength of our realist review was the input of stakeholder groups, particularly the lived experience group, which was widely consulted, and a research team with subject matter expertise to develop and validate the programme theory and CMOCs. Other strengths of our review included the use of Google Alerts to identify more recently published papers (2022–2024) and iterative search techniques to locate other, relevant documents. References from review papers and RCT studies were used to locate kinship and sibling papers. Another key strength of this programme of research is the current collection of primary data to further refine the review’s programme theory and CMOCs.^20^

A key limitation of this study is the lack of grey literature. As outlined in our protocol,^20^ we had intended to include grey literature in this review, however due to the volume of evidence obtained through formal literature searching, this was not possible.

Many intervention studies were RCTs. In most instances, outcomes were reported at the conclusion of the studies with lack of follow-up to determine sustainability of interventions. The long-term effectiveness of the interventions reported in this review, therefore, is not known.

## Conclusion

Our realist review identified twelve CMOCs and an accompanying theory of individuals’ recovery and weight management—intertwined goals for individuals with SMI and weight gain from anti-psychotic medications. Successful interventions are characterized by proactive interventions with holistic comprehensive assessment and appropriate behaviour change therapies to promote individual engagement in community-based weight management programmes. The interventions are underpinned by trusting and honest therapeutic relationships with practitioners with a de-stigmatising approach and open sharing of weight gain health risks.

Necessary support includes family/carers and friends, practitioners, train from ed facilitators, peer group participants and peer mentors. Weight management programmes need to be flexible, but promising models of community-based services for this population feature collaboration and cooperation across secondary and primary services with a primary contact for support. The programme theory with its 12 CMOCs provide testable explanations for what works, for whom, when, how and why. This knowledge can be used to inform practice and research in the future.

## Data Availability

All data produced in the present study are available upon reasonable request to the authors

## Acknowledgements

This publication presents independent research commissioned by the National Institute for Health and Care Research (NIHR: HSDR grant number - 131871). The views and opinions expressed by authors in this publication are those of the authors and do not necessarily reflect those of the NIHR or the Department of Health and Social Care. RU is supported by the NIHR Oxford Health Biomedical Research Centre. AA is funded by the Medical Research Council (grant MC_UU_00006/6) and the NIHR Cambridge Biomedical Research Centre. We thank Claire Duddy for conducting the searches for this review.

