## Supplemental Files for "REalist Synthesis Of non-pharmacologicaL interVEntions for antipsychotic-induced weight gain (RESOLVE) in people living with severe mental illness"

**Authors:** Maura MacPhee (University of British Columbia), Jo Howe (Aston University)*, Hafsah Habib (Aston University), Emilia Piwowarczyk (University of British Columbia), Geoff Wong (University of Oxford), Amy Ahern (University of Cambridge), Gurkiran Birdi (Aston University), Suzanne Higgs (University of Birmingham), Sheri Oduola (University of East Anglia), Alex Kenny (McPin Foundation), Annabel Walsh (McPin Foundation), Rachel Upthegrove (University of Birmingham, Early Intervention Service, Birmingham Women's and Children's NHS Foundation Trust), Katherine Allen (Birmingham and Solihull NHS Mental Health Foundation Trust), Max Carlish (Birmingham and Solihull NHS Mental Health Foundation Trust), Justine Lovell (Birmingham and Solihull NHS Mental Health Foundation Trust), Ian Maidment (Aston University)

### Supplementary File 1: RESOLVE search strategies

#### Main database search

##### MEDLINE

Host: Ovid

Data parameters: Ovid MEDLINE® ALL

Date range searched: 1946 to present (Daily update)

Date searched: 15/02/2022

Searcher: CD

Hits n=255

| | | |
|---|---|---|
| 1 | (serious mental illness* or serious mental disorder*).ti,ab,kw. | 4826 |
| 2 | (severe mental illness* or severe mental disorder*).ti,ab,kw. | 6829 |
| 3 | SMI.ti,ab,kw. | 5820 |
| 4 | (schizophren* or schizoaffective or psychosis or psychotic or bipolar disorder* or personality disorder*).ti,ab,kw. | 215735 |
| 5 | exp *"bipolar and related disorders"/ or exp *"schizophrenia spectrum and other psychotic disorders"/ or exp *"personality disorders"/ | 184663 |
| 6 | or/1-5 | 275737 |
| 7 | (antipsychotic* or anti-psychotic* or psychotropic* or neuroleptic*).ti,ab,kw. | 76665 |
| 8 | (clozapine or olanzapine or quetiapine or risperidone or aripiprazole or haloperidol).ti,ab,kw. | 43723 |
| 9 | exp *Antipsychotic Agents/ | 79729 |
| 10 | or/7-9 | 135132 |
| 11 | (overweight or obese or obesity).ti,ab,kw. | 362647 |
| 12 | (weight adj1 (loss* or reduc* or gain* or change* or manag* or control*)).ti,ab,kw. | 189676 |
| 13 | (body mass index or BMI).ti,ab,kw. | 283566 |
| 14 | exp *body weight changes/ or exp *overweight/ | 197201 |
| 15 | *body mass index/ | 23299 |
| 16 | or/11-15 | 680164 |
| 17 | ((non-pharma* or nonpharma*) adj2 (intervention* or program*)).ti,ab,kw. | 6614 |
| 18 | ((life-style or lifestyle or healthy living) adj2 (modification* or change* or improv* or intervention* or program* or educat*)).ti,ab,kw. | 31773 |
| 19 | ((nutrition* or diet or healthy eating) adj2 (modification* or change* or improv* or intervention* or program* or educat*)).ti,ab,kw. | 50867 |
| 20 | ((exercise or physical* activ* or physical* training* or sport) adj2 (modification* or change* or improv* or intervention* or program* or educat*)).ti,ab,kw. | 58196 |
| 21 | exp *Life Style/ | 43666 |
| 22 | *health education/ or exp *health promotion/ | 88576 |
| 23 | exp *Exercise/ | 150806 |
| 24 | exp *Sports/ | 140318 |
| 25 | exp *Diet/ | 146521 |
| 26 | exp *Diet Therapy/ | 33186 |
| 27 | ((psycho-educat* or psychoeducat* or behavio* or cognitive or motivation*) adj2 (intervention* or program* or educat* or therap* or counsel*)).ti,ab,kw. | 72092 |
| 28 | exp *Cognitive Behavioral Therapy/ | 24775 |
| 29 | *counseling/ or *directive counseling/ or *motivational interviewing/ | 20109 |
| 30 | or/17-29 | 670437 |
| 31 | 6 and 10 and 16 and 30 | 274 |
| 32 | limit 31 to english language | 255 |

#### Embase

Host: Ovid

Data parameters: Embase 1974 to present

Date range searched: 1974 to present (Daily update)

Date searched: 15/02/2022

Searcher: CD

Hits: n=442

| | | |
|---|---|---|
| 1 | (serious mental illness* or serious mental disorder*).ti,ab,kw. | 5903 |
| 2 | (severe mental illness* or severe mental disorder*).ti,ab,kw. | 9085 |
| 3 | SMI.ti,ab,kw. | 8626 |
| 4 | (schizophren* or schizoaffective or psychosis or psychotic or bipolar disorder* or personality disorder*).ti,ab,kw. | 290260 |
| 5 | exp *bipolar disorder/ or exp *schizophrenia/ or exp *psychosis/ or exp *personality disorder/ | 233421 |
| 6 | or/1-5 | 349112 |
| 7 | (antipsychotic* or anti-psychotic* or psychotropic* or neuroleptic*).ti,ab,kw. | 112006 |
| 8 | (clozapine or olanzapine or quetiapine or risperidone or aripiprazole or haloperidol).ti,ab,kw. | 61456 |
| 9 | exp *neuroleptic agent/ | 130170 |
| 10 | or/7-9 | 202471 |
| 11 | (overweight or obese or obesity).ti,ab,kw. | 535828 |
| 12 | (weight adj1 (loss* or reduc* or gain* or change* or manag* or control*)).ti,ab,kw. | 283921 |
| 13 | (body mass index or BMI).ti,ab,kw. | 512301 |
| 14 | exp *obesity/ | 265482 |
| 15 | exp *body weight change/ or exp *body weight control/ | 14276 |
| 16 | *body mass/ | 36582 |
| 17 | or/11-16 | 1074014 |
| 18 | ((non-pharma* or nonpharma*) adj2 (intervention* or program*)).ti,ab,kw. | 8553 |
| 19 | ((life-style or lifestyle or healthy living) adj2 (modification* or change* or improv* or intervention* or program* or educat*)).ti,ab,kw. | 47125 |
| 20 | ((nutrition* or diet or healthy eating) adj2 (modification* or change* or improv* or intervention* or program* or educat*)).ti,ab,kw. | 68998 |
| 21 | ((exercise or physical* activ* or physical* training* or sport) adj2 (modification* or change* or improv* or intervention* or program* or educat*)).ti,ab,kw. | 80151 |
| 22 | exp *lifestyle/ or exp *lifestyle modification/ | 33552 |
| 23 | exp *health education/ or *diabetes education/ or *nutrition education/ or *patient education/ or *psychoeducation/ | 118646 |
| 24 | *health promotion/ | 38698 |
| 25 | exp *exercise/ | 159086 |
| 26 | exp *sport/ | 82162 |
| 27 | exp *diet/ | 114237 |
| 28 | exp *diet therapy/ | 100952 |
| 29 | ((psycho-educat* or psychoeducat* or behavio* or cognitive or motivation*) adj2 (intervention* or program* or educat* or therap* or counsel*)).ti,ab,kw. | 93939 |
| 30 | exp *cognitive behavioral therapy/ | 7045 |
| 31 | exp *counseling/ | 39906 |
| 32 | or/18-31 | 815018 |
| 33 | 6 and 10 and 17 and 32 | 473 |

| | | |
|---|---|---|
| 34 | limit 33 to english language | 442 |

#### PsycINFO

Host: Ovid

Data parameters: PsycINFO 1806 to present

Date range searched: 1806 to present (Weekly update)

Date searched: 15/02/2022

Searcher: CD

Hits: n=184

| | | |
|---|---|---|
| 1 | (serious mental illness* or serious mental disorder*).ti,ab. | 5557 |
| 2 | (severe mental illness* or severe mental disorder*).ti,ab. | 6930 |
| 3 | SMI.ti,ab. | 2521 |
| 4 | (schizophren* or schizoaffective or psychosis or psychotic or bipolar disorder* or personality disorder*).ti,ab. | 216062 |
| 5 | exp *psychosis/ or exp *acute psychosis/ or exp *chronic psychosis/ or exp *"paranoia (psychosis)"/ or exp *schizophrenia/ or exp *personality disorders/ or exp *bipolar disorder/ | 159662 |
| 6 | or/1-5 | 242118 |
| 7 | (antipsychotic* or anti-psychotic* or psychotropic* or neuroleptic*).ti,ab. | 51212 |
| 8 | (clozapine or olanzapine or quetiapine or risperidone or aripiprazole or haloperidol).ti,ab. | 25873 |
| 9 | exp *neuroleptic drugs/ | 28709 |
| 10 | or/7-9 | 64257 |
| 11 | (overweight or obese or obesity).ti,ab. | 46237 |
| 12 | (weight adj1 (loss* or reduc* or gain* or change* or manag* or control*)).ti,ab. | 27676 |
| 13 | (body mass index or BMI).ti,ab. | 30196 |
| 14 | exp *body weight/ or *body mass index/ | 50441 |
| 15 | or/11-14 | 91976 |
| 16 | ((non-pharma* or nonpharma*) adj2 (intervention* or program*)).ti,ab. | 1830 |
| 17 | ((life-style or lifestyle or healthy living) adj2 (modification* or change* or improv* or intervention* or program* or educat*)).ti,ab. | 6264 |
| 18 | ((nutrition* or diet or healthy eating) adj2 (modification* or change* or improv* or intervention* or program* or educat*)).ti,ab. | 6898 |
| 19 | ((exercise or physical* activ* or physical* training* or sport) adj2 (modification* or change* or improv* or intervention* or program* or educat*)).ti,ab. | 13923 |
| 20 | exp *lifestyle/ | 9196 |
| 21 | *health education/ | 11634 |
| 22 | *health promotion/ | 22253 |
| 23 | exp *exercise/ | 23890 |
| 24 | exp *sports/ | 31753 |
| 25 | *diets/ or *dietary restraint/ | 11622 |
| 26 | ((psycho-educat* or psychoeducat* or behavio* or cognitive or motivation*) adj2 (intervention* or program* or educat* or therap* or counsel*)).ti,ab. | 83483 |
| 27 | *cognitive behavior therapy/ | 18858 |
| 28 | *counseling/ or *group counseling/ or *life coaching/ | 24766 |
| 29 | *motivational interviewing/ | 2139 |

| | | |
|---|---|---|
| 30 | or/16-29 | 224862 |
| 31 | 6 and 10 and 15 and 30 | 201 |
| 32 | limit 31 to english language | 184 |

#### CINAHL (Cumulative Index to Nursing and Allied Health Literature)

Host: EbscoHOST

Data parameters: CINAHL 1981 onwards

Date range searched: 1981 to present (Update unknown)

Date searched: 15/02/2022

Searcher: CD

Hits: n=133

| | | |
|---|---|---|
| S34 | S6 AND S10 AND S17 AND S32 | 133 |
| S33 | S6 AND S10 AND S17 AND S32 | 134 |
| S32 | S18 OR S19 OR S20 OR S21 OR S22 OR S23 OR S24 OR S25 OR S26 OR S27 OR S28 OR S29 OR S30 OR S31 | 511,102 |
| S31 | (MM "Counseling") OR (MM "Motivational Interviewing") OR (MM "Peer Counseling") | 17,741 |
| S30 | (MM "Cognitive Therapy") | 11,664 |
| S29 | TI ( ((psycho-educat* or psychoeducat* or behavio* or cognitive or motivation*) N2 (intervention* or program* or educat* or therap* or counsel*)) ) OR AB ( ((psycho-educat* or psychoeducat* or behavio* or cognitive or motivation*) N2 (intervention* or program* or educat* or therap* or counsel*)) ) | 46,139 |
| S28 | (MM "Diet Therapy+") | 19,599 |
| S27 | (MM "Diet+") | 74,008 |
| S26 | (MM "Sports+") | 63,458 |
| S25 | (MM "Exercise+") | 76,607 |
| S24 | (MM "Health Promotion") | 44,213 |
| S23 | (MM "Health Education+") | 63,536 |
| S22 | (MM "Life Style+") | 130,614 |
| S21 | TI ( ((exercise or physical* activ* or "physical* training*" or sport) N2 (modification* or change* or improv* or intervention* or program* or educat*)) ) OR AB ( ((exercise or physical* activ* or "physical* training*" or sport) N2 (modification* or change* or improv* or intervention* or program* or educat*)) ) | 39,742 |
| S20 | TI ( ((nutrition* or diet or "healthy eating") N2 (modification* or change* or improv* or intervention* or program* or educat*)) ) OR AB ( ((nutrition* or diet or "healthy eating") N2 (modification* or change* or improv* or intervention* or program* or educat*)) ) | 27,276 |

| | | |
|---|---|---|
| S19 | TI ( ((life-style or lifestyle or "healthy living") N2 (modification* or change* or improv* or intervention* or program* or educat*)) ) OR AB ( ((life-style or lifestyle or "healthy living") N2 (modification* or change* or improv* or intervention* or program* or educat*)) ) | 17,133 |
| S18 | TI ( ((non-pharma* or nonpharma*) N2 (intervention* or program*)) ) OR AB ( ((non-pharma* or nonpharma*) N2 (intervention* or program*)) ) | 3,498 |
| S17 | S11 OR S12 OR S13 OR S14 OR S15 OR S16 | 228,745 |
| S16 | (MM "Body Mass Index") | 13,653 |
| S15 | (MM "Obesity") OR (MM "Obesity, Morbid") | 60,201 |
| S14 | (MM "Body Weight Changes") | 544 |
| S13 | TI ( ("body mass index" OR BMI) ) OR AB ( ("body mass index" OR BMI) ) | 101,721 |
| S12 | TI ( (weight N1 (loss* or reduc* or gain* or change* or manag* or control*)) ) OR AB ( (weight N1 (loss* or reduc* or gain* or change* or manag* or control*)) ) | 58,431 |
| S11 | TI ( overweight OR obese OR obesity ) OR AB ( overweight OR obese OR obesity ) | 122,580 |
| S10 | S7 OR S8 OR S9 | 27,505 |
| S9 | (MM "Antipsychotic Agents+") | 14,094 |
| S8 | TI ( clozapine or olanzapine or quetiapine or risperidone or aripiprazole or haloperidol ) OR AB ( clozapine or olanzapine or quetiapine or risperidone or aripiprazole or haloperidol ) | 8,776 |
| S7 | TI ( antipsychotic* OR anti-psychotic* OR psychotropic* OR neuroleptic* ) OR AB ( antipsychotic* OR anti-psychotic* OR psychotropic* OR neuroleptic* ) | 19,096 |
| S6 | S1 OR S2 OR S3 OR S4 OR S5 | 142,246 |
| S5 | (MM "Psychotic Disorders+") OR (MM "Personality Disorders+") | 117,596 |
| S4 | TI ( schizophren* OR schizoaffective OR psychosis OR psychotic OR "bipolar disorder*" OR "personality disorder*" ) OR AB ( schizophren* OR schizoaffective OR psychosis OR psychotic OR "bipolar disorder*" OR "personality disorder*" ) | 57,483 |
| S3 | TI SMI OR AB SMI | 2,256 |
| S2 | TI ( "severe mental illness*" OR "severe mental disorder*" ) OR AB ( "severe mental illness*" OR "severe mental disorder*" ) | 3,710 |
| S1 | TI ( "serious mental illness*" OR "serious mental disorder*" ) OR AB ( "serious mental illness*" OR "serious mental disorder*" ) | 3,399 |

#### Cochrane Library

Host: Cochrane Library

Data parameters: CDSR, Protocols, CENTRAL (trials), Editorials, Special Collections, Clinical Answers

Date range searched: No limit

Date searched: 15/02/2022

Searcher: CD

Hits: n=19

| | | |
|---|---|---|
| #1 | ("serious mental illness*" or "serious mental disorder*"):ti,ab,kw | 740 |
| #2 | ("severe mental illness*" or "severe mental disorder*"):ti,ab,kw | 1009 |
| #3 | (SMI):ti,ab,kw | 843 |
| #4 | (schizophren* or schizoaffective or psychosis or psychotic or "bipolar disorder*" or "personality disorder*"):ti,ab,kw | 30312 |
| #5 | MeSH descriptor: [Bipolar and Related Disorders] explode all trees | 2824 |
| #6 | MeSH descriptor: [Schizophrenia Spectrum and Other Psychotic Disorders] explode all trees | 9664 |

| | | |
|---|---|---|
| #7 | MeSH descriptor: [Personality Disorders] explode all trees | 1460 |
| #8 | #1 OR #2 OR #3 OR #4 OR #5 OR #6 OR #7 | 31896 |
| #9 | *(antipsychotic* or anti-psychotic* or psychotropic* or neuroleptic*):ti,ab,kw* | 14117 |
| #10 | (clozapine or olanzapine or quetiapine or risperidone or aripiprazole or haloperidol):ti,ab,kw | 10968 |
| #11 | MeSH descriptor: [Antipsychotic Agents] explode all trees | 4782 |
| #12 | #9 OR #10 OR #11 | 19319 |
| #13 | (overweight or obese or obesity):ti,ab,kw | 50526 |
| #14 | (weight near/1 (loss* or reduc* or gain* or change* or manag* or control*)):ti,ab,kw | 40770 |
| #15 | ("body mass index" or BMI):ti,ab,kw | 69963 |
| #16 | #12 OR #13 OR #14 OR #15 | 132350 |
| #17 | ((non-pharma* or nonpharma*) near/2 (intervention* or program*)):ti,ab,kw | 1809 |
| #18 | ((life-style or lifestyle or "healthy living") near/2 (modification* or change* or improv* or intervention* or program* or educat*)):ti,ab,kw | 12405 |
| #19 | ((nutrition* or diet or "healthy eating") near/2 (modification* or change* or improv* or intervention* or program* or educat*)):ti,ab,kw | 14913 |
| #20 | ((exercise or "physical* activ*" or "physical* training*" or sport) near/2 (modification* or change* or improv* or intervention* or program* or educat*)):ti,ab,kw | 27997 |
| #21 | MeSH descriptor: [Life Style] explode all trees | 6119 |
| #22 | MeSH descriptor: [Health Promotion] explode all trees | 7041 |
| #23 | MeSH descriptor: [Health Education] this term only | 4139 |
| #24 | MeSH descriptor: [Exercise] explode all trees | 27529 |
| #25 | MeSH descriptor: [Sports] explode all trees | 16826 |
| #26 | MeSH descriptor: [Diet] explode all trees | 19890 |
| #27 | MeSH descriptor: [Diet Therapy] explode all trees | 6425 |
| #28 | ((psycho-educat* or psychoeducat* or behavio* or cognitive or motivation*) near/2 (intervention* or program* or educat* or therap* or counsel*)):ti,ab,kw | 48897 |
| #29 | MeSH descriptor: [Cognitive Behavioral Therapy] explode all trees | 10043 |
| #30 | MeSH descriptor: [Counseling] this term only | 4501 |
| #31 | MeSH descriptor: [Directive Counseling] this term only | 413 |
| #32 | MeSH descriptor: [Motivational Interviewing] this term only | 971 |
| #33 | #17 OR #18 OR #19 OR #20 OR #21 OR #22 OR #23 OR #24 OR #25 OR #26 OR #27 OR #28 OR #29 OR #30 OR #31 OR #32 | 145851 |
| #34 | #8 AND #12 AND #16 AND #33 | 787 |

Of which 18 Cochrane reviews
1 Cochrane protocol
768 trials (not exported)

#### Scopus

Host: Scopus.com

Data parameters: n/a

Date range searched: No limit

Date searched: 16/02/2022

Searcher: CD

Hits: n=224

| | |
|---|---|
| Full string | ( ( TITLE-ABS ( "serious mental illness*" OR "serious mental disorder*" OR "severe mental illness*" OR "severe mental disorder*" OR smi OR schizophren* OR schizoaffective OR psychosis OR psychotic OR "bipolar disorder*" OR "personality disorder*" ) ) ) AND ( ( TITLE-ABS ( antipsychotic OR anti-psychotic* OR psychotropic* OR neuroleptic* OR clozapine OR olanzapine OR quetiapine OR risperidone OR aripiprazole OR haloperidol ) ) ) AND ( TITLE-ABS ( ( overweight OR obese OR obesity OR ( weight W/1 ( loss* OR reduc* OR gain* OR change* OR manag* OR control* ) ) OR "body mass index" OR bmi ) ) ) AND ( TITLE-ABS ( ( ( non-pharma* OR nonpharma* ) W/2 ( intervention* OR program* ) ) OR ( ( life-style OR lifestyle OR "healthy living" OR nutrition* OR diet OR "healthy eating" OR exercise OR "physical* activ*" OR "physical* training*" OR sport ) W/2 ( modification* OR change* OR improv* OR intervention* OR program* OR educat* ) ) OR ( ( psycho-educat* OR psychoeducat* OR behavio* OR cognitive OR motivation* ) W/2 ( intervention* OR program* OR educat* OR therap* OR counsel* ) ) ) ) |
| | Limited to English only |
| Notes | Limited to title/abstract because including keywords quadrupled hits and became overly sensitive |

#### Web of Science (Core)

Host: Web of Science (Clarivate Analytics)

Data parameters: SCIE, SSCI, SHCI, ESCI, CPCI, BKCI indexes

Date range searched: Unknown

Date searched: 16/02/2022

Searcher: CD

Hits: n=204

| | |
|---|---|
| 1 | TI=(("Serious mental illness*" OR "serious mental disorder*" OR "severe mental illness*" OR "severe mental disorder*" OR SMI OR schizophren* OR schizoaffective OR psychosis OR psychotic OR "bipolar disorder*" OR "personality disorder*")) OR AB=(("Serious mental illness*" OR "serious mental disorder*" OR "severe mental illness*" OR "severe mental disorder*" OR SMI OR schizophren* OR schizoaffective OR psychosis OR psychotic OR "bipolar disorder*" OR "personality disorder*")) |
| 2 | TI=(antipsychotic* OR anti-psychotic* OR psychotropic* OR neuroleptic* OR clozapine OR olanzapine OR quetiapine OR risperidone OR aripiprazole OR haloperidol) OR AB=(antipsychotic* OR anti-psychotic* OR psychotropic* OR neuroleptic* OR clozapine OR olanzapine OR quetiapine OR risperidone OR aripiprazole OR haloperidol) |
| 3 | TI=(overweight OR obese OR obesity OR (weight near/1 (loss* OR reduc* OR gain* OR change* OR manag* OR control* )) OR "body mass index" OR bmi ) OR AB=(overweight OR obese OR obesity OR (weight near/1 (loss* OR reduc* OR gain* OR change* OR manag* OR control* )) OR "body mass index" OR bmi ) |

| | |
|---|---|
| 4 | TI=(((non-pharma* OR nonpharma*) near/2 ( intervention* OR program* )) OR (( life-style OR lifestyle OR "healthy living" OR nutrition* OR diet OR "healthy eating" OR exercise OR "physical* activ*" OR "physical* training*" OR sport ) near/2 (modification* OR change* OR improv* OR intervention* OR program* OR educat* )) OR (( psycho-educat* OR psychoeducat* OR behavio* OR cognitive OR motivation* ) near/2 ( intervention* OR program* OR educat* OR therap* OR counsel* ))) OR AB=(((non-pharma* OR nonpharma*) near/2 ( intervention* OR program* )) OR (( life-style OR lifestyle OR "healthy living" OR nutrition* OR diet OR "healthy eating" OR exercise OR "physical* activ*" OR "physical* training*" OR sport ) near/2 (modification* OR change* OR improv* OR intervention* OR program* OR educat* )) OR (( psycho-educat* OR psychoeducat* OR behavio* OR cognitive OR motivation* ) near/2 ( intervention* OR program* OR educat* OR therap* OR counsel* ))) |
| 5 | 1 AND 2 AND 3 AND 4 |
| 6 | Languages: English |
| 7 | |
| 8 | |
| 9 | |
| 10 | noft(((non-pharma* OR nonpharma*) N/2 ( intervention* OR program* )) OR (( life-style OR lifestyle OR "healthy living" OR nutrition* OR diet OR "healthy eating" OR exercise OR "physical* activ*" OR "physical* training*" OR sport ) N/2 (modification* OR change* OR improv* OR intervention* OR program* OR educat* )) OR (( psycho-educat* OR psychoeducat* OR behavio* OR cognitive OR motivation* ) N/2 ( intervention* OR program* OR educat* OR therap* OR counsel* ))) |
| 11 | |
| 12 | |
| 13 | |
| 14 | |
| 15 | |
| Note | As for Scopus limited to TI/AB as TS was very sensitive |

#### Sociological Abstracts

Host: Proquest

Data parameters: 1952 to present (update unknown)

Date range searched: 1952 to present

Date searched: 13/01/2022

Searcher: CD

Hits: n=0

| | |
|---|---|
| Full search string | (noft("serious mental illness*" OR "serious mental disorder*" OR "severe mental illness*" OR "severe mental disorder*" OR SMI) OR noft(schizophren* OR schizoaffective OR psychosis OR psychotic OR "bipolar disorder*" OR "personality disorder*")) AND noft(antipsychotic* OR anti-psychotic* OR psychotropic* OR neuroleptic* OR clozapine OR olanzapine OR quetiapine OR risperidone OR aripiprazole OR haloperidol) AND noft(overweight OR obese OR obesity OR (weight N/1 (loss* OR reduc* OR gain* OR change* OR manag* OR control* )) OR "body mass index" OR bmi ) AND noft(((non-pharma* OR nonpharma*) NEAR/2 (intervention* OR program*)) OR ((life-style OR lifestyle OR "healthy living" OR nutrition* OR diet OR "healthy eating" OR exercise OR "physical* activ*" OR "physical* training*" OR sport) NEAR/2 (modification* OR change* OR improv* OR intervention* OR program* OR educat*)) OR ((psycho-educat* OR psychoeducat* OR behavio* OR cognitive OR motivation*) NEAR/2 (intervention* OR program* OR educat* OR therap* OR counsel*))) |
| | (Limited to English language) |

### Supplementary File 2: Documents included

This table provides full details of the documents included in the review

*Table 1: RESOLVE Table of Included Documents*

| Title | Author | Year | Country | Source | Population studied | Manuscript type | Summary of findings |
|---|---|---|---|---|---|---|---|
| Weight stigma interventions as future avenues for stigma resistance: comment on Dubreucq et al. | Alliende & Mittal | 2023 | US | Google scholar alert | N/A | Commentary | N/A |
| Psychotic Disorders, Eating Habits, and Physical Activity: Who Is Ready for Lifestyle Changes? | Archie et al | 2007 | Canada | Medline | DSM-IV diagnosis of schizophrenia, schizoaffective disorder, delusional disorder, bipolar disorder, or depression with psychotic features or a first episode of psychosis | Cross-sectional survey design | A total of 101 participants (64 men) (mean±SD age 35±11 years) were taking antipsychotic medications. Seventy-one percent had schizophrenia spectrum disorders, and 15% had affective psychosis. The prevalence of patients identified as being ready for change was higher than expected: 68% for eating habits and 54% for physical activity. Participants who were ready to change eating habits were also ready to change physical activity habits (p<.04). Stages of change for eating habits were associated with body mass index (p<.004), whereas stages of change for physical activity were associated with self-reported vigorous (p<.001) and moderate (p<.005) physical activity but not mild physical activity. |

| | | | | | | | |
|---|---|---|---|---|---|---|---|
| A Qualitative Study of Social Facilitators and Barriers to Health Behavior Change Among Persons with Serious Mental Illness | Aschbrenner et al | 2013 | US | Citation searching from Lee et al, 2022 | Serious mental illness (schizophrenia, schizoaffective disorder, major depression, or bipolar disorder) | Qualitative | Thematic analysis of focus group discussions revealed that emotional, practical, and mutual support from family members and significant others were social facilitators to health behavior change, while unhealthy social environments was a barrier. Participants in the ''high'' achiever group reported more mutual support for health behavior change than participants in the ''low'' achiever group. |
| Peer health coaching for overweight and obese individuals with serious mental illness: intervention development and initial feasibility study | Aschbrenner et al | 2015 | US | Citation searching from Lee et al, 2022 | axis I diagnosis of major depression, bipolar disorder, schizoaffective disorder, or schizophrenia | Pilot feasability study Mixed methods inc qualitative interviews | There was no overall significant change in mean weight; however, over half (56 %) of participants lost weight by the end of the intervention with mean weight loss 2.7± 2.1 kg. Participants reported high satisfaction and perceived benefits from the program. Qualitative interviews with key stakeholders indicated that the intervention was implemented as planned. |
| Feasibility Study of Increasing Social Support to Enhance a Healthy Lifestyle Intervention for Individuals with Serious Mental Illness | Aschbrenner et al | 2016 | US | Cluster search on author | serious mental illness, defined as an Axis I diagnosis of major depression, bipolar disorder, schizoaffective disorder, or schizophrenia; | Feasability study | The majority of participants (57%) nominated a friend, followed by adult child-parent pairs (28%) and sibling pairs (14%) to participate as support partners in the study. All participant-partner dyads (100%) completed 12 sessions within 16 weeks. Participants reported high satisfaction and perceived benefits from the program. Recommend modifications by the dyads included more interactive sessions, a combination of group and dyadic sessions, and hands-on cooking classes. |

| | | | | | | | |
|---|---|---|---|---|---|---|---|
| Healthy Eating in Persons with Serious Mental Illnesses: Understanding and Barriers | Barre et al | 2011 | US | Citation searching from Lee et al, 2022 | a diagnosis of schizophrenia, schizoaffective disorder, bipolar disorder and/or major depression | Qualitative | Most participants described healthy eating as consuming fruits and vegetables, using low fat cooking methods, and limiting sweets, sodas, fast food, and/or junk food. Internal barriers to nutritional change included negative perceptions of healthy eating, the decreased taste and satiation of healthy foods, difficulty changing familiar eating habits, eating for comfort, and the prioritization of mental health. External barriers were the reduced availability and inconvenience of healthy foods, social pressures, and psychiatric medication side effects. |

| | | | | | | | |
|---|---|---|---|---|---|---|---|
| Pragmatic Replication Trial of Health Promotion Coaching for Obesity in Serious Mental Illness and Maintenance of Outcomes | Bartels et al | 2015 | US | Citation searching from Lee et al, 2022 | Serious mental illness, defined as an axis I diagnosis of major depression, bipolar disorder, schizoaffective disorder, or schizophrenia (based on the Structured Clinical Interview for DSM-IV Axis I Disorders, Patient Edition [SCID; 16]) and persistent impairment in multiple areas of functioning (e.g., work, school, self-care) | RCT (Pragmatic replication trial) | Participants (N=210) were ethnically diverse (46% were nonwhite), with a mean baseline BMI of 36.8 (SD=8.2). At 12 months, the In SHAPE group (N=104) had greater reduction in weight and improved fitness compared with the fitness club membership only group (N=106). Primary outcomes were maintained at 18 months. Approximately half of the In SHAPE group (51% at 12 months and 46% at 18 months) achieved clinically significant cardiovascular risk reduction (a weight loss $5% or an increase of .50 meters on the 6-minute walk test). |

| | | | | | | | |
|---|---|---|---|---|---|---|---|
| Implementation of a Lifestyle Intervention for People With Serious Mental Illness in State-Funded Mental Health Centers | Bartels et al | 2018 | US | Cluster search on author | serious mental illness, defined as a chart axis I DSM-IV primary psychiatric diagnosis of schizophrenia, schizoaffective disorder, bipolar disorder, or major depression, with moderate impairment in multiple areas of functioning; | Quasi-experimental observational | Participats in the two In SHAPE sites (N=63 participants)<br>lost significantly more weight (p=.003) and showed<br>greater improvement in fitness (p=.011) compared with participants<br>at the two usual care control sites (N=59 participants).<br>At six months, nearly half (49%) of In SHAPE participants and<br>at 12 months more than half (60%) of In SHAPE participants<br>showed clinically significant cardiovascular risk reduction<br>defined as $5% weight loss or improved fitness (.50 m<br>[164 feet] increase on the six-minute walk test). The difference<br>between the In SHAPE and control groups was not statistically<br>significant. |
| Using pedometers to Document Physical Activity in Patients with Schizophrenia Spectrum Disorders A Feasibility Study | Beebe & Harris | 2012 | US | Cluster search on author | chart diagnosis of schizophrenia, schizoaffective disorder, schizophreniform disorder, or any subtype of these, according to the criteria in the Diagnostic and Statistical Manual of Mental Disorders (APA, 2000); | Feasability study | All participants<br>completed the 1-week data collection period. Twenty-one<br>(87.5%) participants wore their pedometer at least 6 of the 7<br>days. Diffi culties with the pedometers were few and minor.<br>These preliminary fi ndings indicate that a majority of individuals<br>with SSDs are willing to wear pedometers for activity<br>monitoring and can do so with few problems. Possible barriers<br>to the use of extended-wear pedometers in this group<br>include cognitive and memory defi cits. |

| | | | | | | | |
|---|---|---|---|---|---|---|---|
| Feasibility of the Walk, Address, Learn and Cue (WALC) Intervention for Schizophrenia Spectrum Disorders | Beebe & Smith | 2010 | US | Cluster search on author | a chart diagnosis of schizoaffective disorder or schizophrenia, any subtype, according to the criteria described in the Diagnostic and Statistical Manual for Mental Disorders (DSM-IV, American Psychiatric Association, 2000), | Feasability study | The intervention was feasible and acceptable to participants; approximately two-thirds of all groups were attended and nearly half of participants attended at least 75% of groups. |
| Effect of a Motivational Group Intervention on Exercise Self-Efficacy and Outcome Expectations for Exercise in Schizophrenia Spectrum Disorders | Beebe et al | 2010 | US | Citation from Speyer | a chart diagnosis of schizoaffective disorder, schizophrenia (any subtype), or schizophreniform disorder, according to the criteria described in the Diagnostic and Statistical Manual for Mental Disorders, text revision (American Psychiatric Association, 2000); | RCT | N = 97, 46% female, 43% African American, average age 46.9 years (SD = 2.0). Mean SEE scores were significantly higher in WALC-S participants after intervention, F(1, 95) = 5.92, p = .0168, however, mean OEES scores were significantly higher in control participants after intervention, F(1, 95) = 5.76, p = .0183. |

| | | | | | | | |
|---|---|---|---|---|---|---|---|
| Factors Associated with Weight Intervention Participation among People with Serious Mental Illness | Bennett et al | 2018 | US | Cluster search Bennett | Serious mental illness | RCT | SE = self-efficacy<br><br>SE significantly correlated with intervention participation (p<.02). RtC did not predict significantly over and above SE. A linear combination of all measures was significantly related to participation (p<.05). To improve weight intervention participation by individuals with SMI, one direction may be to improve weight loss SE. |
| Preventing the Development of Metabolic Syndrome in People with Psychotic Disorders—Difficult, but Possible: Experiences of Staff Working in Psychosis Outpatient Care in Sweden | Bergqvist et al | 2013 | Sweden | Embase | Staff with at least one year's experience of working in outpatient psychosis care. | Qualitative | The results illustrate that implementation of lifestyle changes among people with psychotic disorders was experienced as difficult, but possible. The greatest obstacles experienced in this work were difficulties due to the reduction of cognitive functions associated with the disease. Guidelines available to staff in order to help them identify and prevent physical health problems in the group were not always followed and the content was not always relevant. Staff further described feelings of uncertainty about having to motivate people to take anti-psychotic medication while simultaneously being aware of the risks of metabolic deviations. Nursing interventions focusing on organising daily routines before conducting a more active prevention of metabolic syndrome, including information and practical support, were experienced as necessary. The importance of healthy eating and physical activity needs to be communicated in such a way that it is adjusted to the person's |

| | | | | | | | |
|---|---|---|---|---|---|---|---|
| | | | | | | | cognitive ability,<br>and should be repeated over time, both verbally and in writing.<br>Such efforts, in combination with empathic and seriously committed<br>community-based social support, were experienced as having<br>the best effect over time. Permanent lifestyle changes were experienced<br>as having to be carried out on the patient's terms and in his<br>or her home environment. |
| "We're all in this together": Peer-specialist Contributions to a Healthy Lifestyle Intervention for People with Serious Mental Illness | Bochicchio et al | 2019 | US | Citation searching from Lee et al, 2022 | Intervention participants, peer specialists, and peers' supervisors<br><br>Intervention participants - diagnosis of serious mental illness | Qualitative (incorporating descriptive statistics) | Peer specialists' disclosure of their own experiences making health behaviors changes was critical for building participants' motivation and ability to try lifestyle changes. Findings can inform peer specialist training and practice standards and facilitate the expansion of peer-delivered interventions to improve the physical health of people with SMI. |
| "Being There" vs "Being Direct:" Perspectives of Persons with Serious Mental Illness on Receiving Support with Physical Health from Peer and Non-Peer Providers | Bochicchio et al | 2021 | US | Cluster search on author | Serious mental illness | Qualitative (incorporating descriptive statistics) | Participants viewed their relationships with peer and non-peer providers positively, but described differences in the approach to practice, power dynamics present, and how they identified with each provider. Participants described peers as process-oriented while non-peer staff as task-oriented, focusing on accomplishing concrete objectives. Each provider sought to boost participants' motivation, but peers built hope by emphasizing the possibility of change, while non-peer providers emphasized the consequences of inaction. Participants related to peer staff through shared experiences, while identifying the importance of having a shared treatment goal with their non-peer provider. |

| | | | | | | | |
|---|---|---|---|---|---|---|---|
| Developing a healthy living intervention for people with early psychosis using the Medical Research Council's guidelines on complex interventions: Phase 1 of the HELPER – InterACT programme | Bradshaw et al | 2012 | UK | Cluster search on author | Early psychosis | Methodological: description of intervention development | The intervention developed comprised eight individual sessions to be delivered by a support time recovery worker over a 12 month period with emphasis on individualised action plans to facilitate participatory exercise and changes in diet. To optimise engagement, choice and self management a booklet and website were developed to provide participants with educational advice, healthy eating recipes and other materials. |
| Treatment response to the RENEW weight loss intervention in schizophrenia: Impact of intervention setting | Brown et al | 2014 | US | Citation searching from Lee et al, 2022 | Serious mental illness | RCT | The intervention group experienced a modest weight loss of 4.8 lbs at 3 months, 4.1 lbs at 6 months and a slight weight gain of 1.5 lbs at 12 months. The control group gained a total of 6.2 lbs at 12 months. However when settings were examined separately the responder sites had a weight loss of 9.4 lbs at 3 months, 10.9 lbs at 6 months and 7 lbs at 12 months. |
| A Pilot Study of the Nutrition and Exercise for Wellness and Recovery (NEW-R): A Weight Loss Program for Individuals With Serious Mental Illnesses | Brown et al | 2015 | US | Cluster search on author | Serious mental illness | Pilot study Pre-post | Participants lost an average of 3 pounds at immediate post-intervention, and lost an average of 10 pounds at the 6 month followup. Participants also demonstrated significant increases in their knowledge about nutrition and physical activity. |
| Main Outcomes of the Peer-Led Healthy Lifestyle Intervention for People with Serious Mental Illness in Supportive Housing | Cabassa et al | 2021 | US | Cluster search on study ID | Serious mental illness | RCT | Participants were predominantly racial/ethnic minorities (81.7%) with a mean baseline weight of 218.8±54.0 pounds and mean Body Mass Index=33.7±7.2. Although a larger proportion of participants in PGLB than in usual care achieved clinically significant changes in study outcomes at 12 and 18 months, none were statistically significant. Outcomes differed by study site: two sites reported no significant differences between PGLB and usual care and one site reported that PGLB significantly outperformed usual care on clinically significant weight loss at |

| | | | | | | | |
|---|---|---|---|---|---|---|---|
| | | | | | | | 18 months and on CVD risk reductions at 6 and 12 months. |
| Reducing weight gain in people with schizophrenia, schizoaffective disorder, and first episode psychosis: describing the process of developing the STructured lifestyle Education for People With SchizophrEnia (STEPWISE) intervention | Carey et al | 2018 | UK | Citation searching from Lee et al, 2022 | Schizophrenia, schizoaffective disorder, and first episode psychosis | Methodological: description of intervention development | This paper reports the process of development, including challenges and how these were addressed. It describes how user input influenced the structure, topics, and approach of the intervention. The outcome of this process was a feasible and acceptable lifestyle intervention to support people with schizophrenia, schizoaffective disorder, or first episode psychosis to manage their weight. This pilot provided opportunities for refinement of the intervention and facilitator training prior to testing in a multi-centre randomised controlled trial. Key findings from the pilot were linked to accessibility, focus, uptake, and retention, which influenced session length, travel arrangements, refreshment, breaks, and supporting tools to incentivise participants. |
| Randomized trial of achieving healthy lifestyles in psychiatric rehabilitation: the ACHIEVE trial | Casagrande et al | 2010 | US | Cluster search on author | Serious mental illness | RCT protocol | N/A |
| Running for your life: A review of physical activity and cardiovascular disease risk reduction in individuals with schizophrenia | Chalfoun et al | 2016 | Canada | Medline | Schizophrenia | Critical literature review | Many non-pharmacological interventions are efficient in reducing cardiovascular disease risk factors when combined with physical activity. Supervised physical activity has been successful in decreasing cardiovascular disease risk, and aerobic interval training appears to provide more benefits by specifically targeting cardiorespiratory fitness levels. |
| Peer Wellness Coaches for Adults With Mental Illness | Cohen et al | 2014 | US | Cluster search on study ID | Serious mental illness | Report | N/A |

| | | | | | | | |
|---|---|---|---|---|---|---|---|
| A Behavioral Weight-Loss Intervention in Persons with Serious Mental Illness | Daumit et al | 2013 | US | Citation searching from Lee et al, 2022 | Serious mental illness | RCT | Of 291 participants who underwent randomization, 58.1% had schizophrenia or a schizoaffective disorder, 22.0% had bipolar disorder, and 12.0% had major depression. At baseline, the mean body-mass index (the weight in kilograms divided by the square of the height in meters) was 36.3, and the mean weight was 102.7 kg (225.9 lb). Data on weight at 18 months were obtained from 279 participants. Weight loss in the intervention group increased progressively over the 18-month study period and differed significantly from the control group at each follow-up visit. At 18 months, the mean between-group difference in weight (change in intervention group minus change in control group) was −3.2 kg (−7.0 lb, P = 0.002); 37.8% of the participants in the intervention group lost 5% or more of their initial weight, as compared with 22.7% of those in the control group (P = 0.009). There were no significant between-group differences in adverse events. |
| A Randomized, Controlled Multisite Study of Behavioral Interventions for Veterans with Mental Illness and Antipsychotic Medication-Associated Obesity | Erickson et al | 2017 | US | Citation from Mucheru | diagnosis of mental illness per DSM-IV | RCT | Participants in both groups lost weight. LB participants had a greater decrease in average waist circumference [F(1,1244) = 11.9, p < 0.001] and percent body fat [F(1,1121) = 4.3, p = 0.038]. Controlling for gender yielded statistically significant changes between groups inBMI [F(1,1246) = 13.9, p < 0.001].Waist circumference and percent body fat decreased for LB women [F(1,1243) = 22.5, p < 0.001 and F(1,1221) = 4.8, p = 0.029, respectively]. The majority of LB participants kept food and activity journals (92%), and average daily calorie intake decreased from2055 to 1650 during the study (p < 0.001). |

| | | | | | | | |
|---|---|---|---|---|---|---|---|
| Physical health—a cluster randomized controlled lifestyle intervention among persons with a psychiatric disability and their staff | Forsberg et al | 2008 | Sweden | Citation searching from Lee et al, 2022 | Psychiatric diagnosis in accordance with DSMIV (e.g. schizophrenia, bipolar disorder, personality disorders, other psychotic disorders and autism spectrum disorders*with no or mild cognitive impairments)<br><br>Staff working with housing support or in supported housing facilities. | RCT | There was a significant reduction in the mean of metabolic syndrome criteria in the intervention group compared with the control group at the follow-up. The participants expressed satisfaction with the programme. |

| | | | | | | | |
|---|---|---|---|---|---|---|---|
| Meanings of participating in a lifestyle programme for persons with psychiatric disabilities | Forsberg et al | 2011 | Sweden | Cluster search on author | diagnosis, in accordance with DSM-IV, of schizophrenia, bipolar disorder, personality disorders, other psychotic disorders and autism spectrum disorders with no or mild cognitive impairments and living in supported living facilities | Qualitative | Meanings of participating in a lifestyle programme include my health can be improved as both the physical effects and the obstacles are considered and the daily life is partially given a changed content in new experiences and by participating in something to take pride in. The meanings of participating together with the staff mean an increased sense of closeness and equality with the staff expressed in changes in relationships and the difference between the two groups being revealed and also in becoming aware of the life situation, an insight into the loss of a healthy life but also hope for the future is expressed. |

| | | | | | | | |
|---|---|---|---|---|---|---|---|
| Mental Health Collaborative Care and its Role in Primary Care Settings | Goodrich et al | 2013 | US | Cluster search on study ID | Serious mental illness | Literature review | Collaborative care models (CCMs) provide a pragmatic strategy to deliver integrated mental health and medical care for persons with mental health conditions served in primary care settings. CCMs are team-based inter- vention to enact system-level redesign by improving patient care through organizational leadership support, provider de- cision support, and clinical information systems, as well as engaging patients in their care through self-management support and linkages to community resources. The model is also a cost-efficient strategy for primary care practices to improve outcomes for a range of mental health conditions across populations and settings. CCMs can help achieve integrated care aims underhealth care reform yet organiza- tional and financial issues may affect adoption into routine primary care. Notably, successful implementation of CCMs in routine care will require alignment of financial incentives to support systems redesign investments, reimbursements for mental health providers, and adaptation across different prac- tice settings and infrastructure to offer all CCM components. |

| | | | | | | | |
|---|---|---|---|---|---|---|---|
| STEPWISE – STructured lifestyle Education for People WIth SchizophrEnia: a study protocol for a randomised controlled trial | Gossage-Worrall et al | 2016 | England | Cluster search on study ID | diagnosis of schizophrenia or schizoaffective disorder (defined by International Classification of Diseases, version 10 (ICD-10) codes F20, F25) or first-episode psychosis (defined as less than 3 years since presentation to the mental health team) using case note review | Protocol | N/A |

| | | | | | | | |
|---|---|---|---|---|---|---|---|
| STructured lifestyle education for people<br>WIth SchizophrEnia (STEPWISE): mixed<br>methods process evaluation of a groupbased<br>lifestyle education programme to support weight loss in people with<br>schizophrenia | Gossage-Worrall et al | 2019 | UK | Citation searching from Lee et al, 2022 | Schizophrenia | Qualitative evaluation of the implementation of the RCT | Training and course materials were available although lacked co-ordination in some trusts. Healthcare professionals gained knowledge and some contemplated changing their practice to reflect the (facilitative) 'style'<br>of delivery. They were often responsible for administrative activities increasing the burden of delivery. Healthcare professionals recognised the need to address antipsychotic-induced weight gain and reported potential value from the intervention (subject to the RCT results). However, some doubted senior management commitment and sustainability post-trial. Service-users found the intervention highly acceptable, especially being in a group of people with similar experiences. Service-users perceived weight loss and lifestyle benefits; however, session attendance varied with 23% (n = 47) attending all group-sessions and 17% (n = 36) attending none. Service-users who lost weight wanted closer monitoring and many healthcare professionals wanted to monitor outcomes (e.g. weight) but it was outside the intervention design. No clinical or cost benefit was demonstrated from the intermediate outcomes (RCT) and any changes in RCT outcomes were not due to the intervention. |
| A 12-Week Weight Reduction Intervention for Overweight Individuals Taking Antipsychotic Medications | Green et al | 2014 | US | Cluster search on author | Taking at least one antipsychotic medication<br>at any consistent dose for a minimum of 30 days at the time<br>they were identified | Feasability trial | Average attendance was 8.6 of 12 sessions. Intent-to-treat analyses of covariance, adjusted for baseline<br>weight, showed significant changes in weight: Mean<br>weight in intervention participants declined from 213.3 to<br>206.6 pounds, while control participants' weight was<br>unchanged. |

| | | | | | | | |
|---|---|---|---|---|---|---|---|
| The STRIDE Weight Loss and Lifestyle Intervention for Individuals Taking Antipsychotic Medications: A Randomized Trial | Green et al | 2015 | US | Citation searching from Lee et al, 2022 | Taking at least one antipsychotic medication at any consistent dose for a minimum of 30 days at the time they were identified | RCT | Participants (56 men, 144 women), mean age = 47.2(*SD* =10.6), were randomized to usual care (n =96) or a 6-month weekly group intervention plus 6 monthly maintenance sessions (n =104). 181 participants (90.5%) completed 6-month, and 170 (85%) completed 12-month assessments, without differential attrition. Participants attended 14.5 of 24 sessions over 6 months. Intent-to-treat analyses found intervention participants lost 4.4 kg more than control participants from baseline to 6 months (95% CI [−6.96 kg, −1.78 kg]), and 2.6 kg more than controls (95% CI −5.14 kg, −0.07 kg] from baseline to 12 months. At 12 months, fasting glucose levels in controls had increased from 106.0 mg/dL to 109.5 mg/dL and decreased in intervention participants, from 106.3 mg/dL to 100.4 mg/dL. No serious adverse events were study-related; medical hospitalizations were reduced in the intervention group (6.7%) compared to controls (18.8%)($\chi 2$= 6.66, $p$ = 0.01). Individuals taking antipsychotic medications can lose weight and improve fasting glucose levels. Increasing reach of the intervention is an important future step. |

| | | | | | | | |
|---|---|---|---|---|---|---|---|
| Effect of lifestyle coaching versus care coordination versus treatment as usual in people with severe mental illness and overweight: Two-years follow-up of the randomized CHANGE trial | Jakobsen et al | 2017 | Denmark | Cluster search on author | diagnosed according to the ICD-10 with schizophrenia (F20), schizoaffective disorder (F25), or persistent delusional disorder (F22)±confirmed at initial assessment by the Schedules for Clinical Assessment in Neuropsychiatry (SCAN) | 2-year follow up of RCT | After two-years the mean 10-year cardiovascular-disease risk was 8.7% (95% confidence interval (CI) 7.6±9.9%) in the CHANGE group, 7.7% (95% CI 6.5±8.9%) in the care coordination group, and 8.9% (95% CI 6.9±9.2%) in the treatment as usual group (P = 0.24). Also, there were no intervention effects for any secondary or exploratory outcomes, including cardiorespiratory fitness, weight, physical activity, diet and smoking. No reported adverse events could be ascribed to the intervention. |
| Understanding Women Veterans' Experiences With and Management of Weight Gain From Medications for Serious Mental Illness: A Qualitative Study | Kreyenbuhl et al | 2019 | US | CINAHL | diagnosed with schizophrenia, schizoaffective disorder, bipolar affective disorder, manic disorder, or psychosis disorder not otherwise specified | Qualitative | We identified 5 themes related to females' experiences with medication-induced weight gain. Female veterans described considerable psychological and physical distress associated with weight gain. However, many expressed a willingness to accept weight gain as a trade-off for medications' therapeutic effects, a theme echoed by prescribers. Both described primarily using reactive rather than proactive or preventative weight management approaches and described the limited effectiveness of reactive approaches. Other contributing factors, including the multiple and uncertain causes of weight gain, uneven quality and quantity of weight loss information, lack of social support, and environmental barriers, add to the difficulty and complexity of their struggles. |

| | | | | | | | |
|---|---|---|---|---|---|---|---|
| Antipsychotic-induced weight gain: exploring the role of psychiatrists in managing patients' physical health – challenges, current options and direction for future care | Lee et al | 2023 | UK | Google scholar alert | Serious mental illness | Commentary | N/A |
| Design of the Lifestyle Interventions for severe mentally ill Outpatients in the Netherlands (LION) trial; a cluster randomised controlled study of a multidimensional web tool intervention to improve cardiometabolic health in patients with severe mental illness | Loojimans et al | 2017 | Netherlands | Cluster search on author | Community-dwelling Severe mental illness patients and Severe mental illness patients living in sheltered facilities. | Methodological: description of intervention development | N/A |
| Functionality appreciation is inversely associated with positive psychotic symptoms in overweight/obese patients with schizophrenia | Mahfoud et al | 2023 | Lebanon | Google scholar alert | Schizophrenia | Cross sectional study | No significant difference was found between overweight/obese and normal-weight patients for all variables, except for weight stigma; a significantly higher weight stigma score was significantly found in overweight/obese compared to normal-weight patient. In the bivariate analysis, higher functionality appreciation was significantly associated with higher positive PANSS scores. The results of the linear regression, taking the positive PANSS score as the dependent variable, showed that higher functionality appreciation (Beta = − 0.52) and higher social support (Beta = − 0.16) were significantly associated with lower positive PANSS scores, whereas having a secondary education level compared to illiteracy (Beta = 7.00) was significantly associated with higher positive PANSS scores. |

| | | | | | | | |
|---|---|---|---|---|---|---|---|
| Weight gain associated with taking psychotropic medication: An integrative review | McCloughen & Foster | 2011 | Australia | Medline | psychoses and mood disorders | Integrative review of research | The review revealed people experiencing psychotropic-induced weight gain perceive it as distressing. It impacts on quality of life and contributes to treatment non-adherence. Weight management and prevention strategies have primarily targeted adults with existing/chronic illness rather than those with firstepisode psychoses and/or drug naiveté. Single and multimodal interventions to prevent or manage weight gain produced comparable, modest results. |
| Barriers and Facilitators of a Healthy Lifestyle Among Persons with Serious and Persistent Mental Illness: Perspectives of Community Mental Health Providers | McKibbin et al | 2014 | US | Cluster search on author | Community mental health providers | Qualitative | Results showed that influences at individual, social, community, and societal levels impact development and maintenance of obesity. Mental health providers desired a collaborative relationship with providers of health promotion program staff. They also believed that frequent, group-based health promotion should include participation incentives for adults with SMI and should occur over durations of at least 6-months to achieve improved health outcomes for this population. |
| Quality of Life Outcomes of Web-Based and In-Person Weight Management for Adults with Serious Mental Illness | Muralidharan et al | 2020 | US | Cluster search on author | chart diagnosis of schizophrenia, schizoaffective disorder, affective psychoses, post-traumatic stress disorder | Secondary analysis of RCT data | Compared to usual care, in-person MOVE was associated with improvements in loneliness (t=−2.76, p=.006) and mental health related quality of life (t=1.99, p=0.048) at six months, and webMOVE was associated with improvements in weight-related self-esteem at six months (t=2.23, p=.026) and mental health-related quality of life at three months (t=2.17, p=0.031) and six months (t=2.38, p=.018). Web-based and in-person weight management led to improvements in health-related quality of life for adults with serious mental illness. |

| | | | | | | | |
|---|---|---|---|---|---|---|---|
| Impact of Online Weight Management with Peer Coaching on Physical Activity Levels of Adults with Serious Mental Illness | Muralidharan et al | 2018 | US | Citation searching from Lee et al, 2022 | serious mental illness diagnosis (schizophrenia spectrum disorders, affective psychoses, or post-traumatic stress disorder) | Secondary analysis of RCT data | Comparing MOVE SMI to usual care, there were significant group differences for total physical activity at 3-months (t=3.06, df=272, p=.002) and 6-months (t=3.12, df=272, p=.002), for walking at 6- months (t=1.99, df=273, p=.048), for moderate physical activity at 6-months (t = 2.12, p =.035), and for vigorous physical activity at 6-months (t = 2.34, p=.020). Comparing WebMOVE to usual care, there was a significant group difference for total physical activity at 6-months (t=2.02, df=272, p=.044), and a trend for a group difference for walking at 6-months (t=1.78, df=273, p=.076). These findings reflected declines in usual care and increases in MOVE SMI or WebMOVE, respectively. |
| Mental Health in Primary Care: Perceptions of Augmented Care for Individuals With Serious Mental Illness | Nover | 2013 | US | Citation searching from Lee et al, 2022 | SMI | Qualitative | Medical service needs included the need to provide input in treatment and to be personally valued. Barriers ranged from doubts about provider qualifications to concerns about medication. Elements of the augmented care program that participants found beneficial were those involving care coordination, social support, and weight management support. |
| Developing a peer-based healthy lifestyle program for people with serious mental illness in supportive housing | O'Hara et al | 2017 | US | Citation searching from Lee et al, 2022 | chart diagnosis of SMI | Pilot study | Participants on average attended 8 of 12 sessions, and reported that services were satisfactory and helpful. Intervention adaptations, ongoing throughout the study, focused on adding peer-specialists as co-facilitators, increasing individualized support and developing strategies to address socioeconomic barriers impacting participants' ability to engage in healthy lifestyle changes. |

| | | | | | | | |
|---|---|---|---|---|---|---|---|
| Physical health problems in people with psychosis: The issue for informal carers | Onwumere et al | 2018 | England | Cluster search on author | Carers of adults with psychotic disorders | Qualitative | Five key themes were identified from the interviews that reflected (1) ubiquity of physical health problems in psychosis, (2) gaps in service provision for those living with mental and physical health problems, (3) carers' role in responding to service gaps, (4) difficult conversations and (5) impact on carer health. |
| Participants' Voices From Within a Healthy Lifestyle Group | Park, Foster & Usher | 2017 | Australia | Citation searching from Lee et al, 2022 | Schizophrenia | Qualitative | Four themes were identified. The findings indicate that benefits of the program were more than physical health improvements and included regular access to a health professional, gaining social relationships, and a sense of belonging. Future recommendations include retaining a group structure in lifestyle interventions to facilitate these additional benefits. |
| Description of a healthy lifestyle intervention for people with serious mental illness taking second-generation antipsychotics | Park, Usher & Foster | 2011 | Australia | Cluster search on author | serious mental illness | Methodological: description of intervention development | N/A |

| | | | | | | | |
|---|---|---|---|---|---|---|---|
| Understanding the problems developing a healthy living programme in patients with serious mental illness: a qualitative study | Pearsall et al | 2014 | Scotland | Citation searching from Lee et al, 2022 | Diagnosis of schizophrenia, schizoaffective or bipolar affective disorder | Qualitative | Thirteen patients were interviewed during the study. Many did not recall receiving an initial invitation letter to the programme. Several believed that there was no necessity to attend as they had already had recent routine health checks by their general practitioner. The patients' current level of mental and physical health was important with symptoms such as depression, anxiety or arthritis affecting interest in the programme. Patients described that they found smoking enjoyable or calming in its effect. Dietary intake was determined by taste or gaining pleasure in eating certain types of food. Several lessons were learnt during this research that may aid future research and practice. Participation seemed to be better if the approach was first made by the patient's own community keyworker. This contact may have provided a greater opportunity to explain the purpose and importance of the programme. Alternative appointments should be considered when certain patients are in better physical and mental health. Healthy living programmes need to be flexible and adaptive to individual patient needs. Assistance from their community worker may help engagement. Simple measures may improve participation and reduce potential barriers. |

| | | | | | | | |
|---|---|---|---|---|---|---|---|
| Collaborative, individualised lifestyle interventions are acceptable to people with first episode psychosis; a qualitative study | Pedley et al | 2018 | England | Cluster search on author | First episode psychosis | Qualitative | Participants valued the collaborative and individualised approach taken by the intervention deliverers, and formed high quality relationships with them. Aspects of the intervention that were positively appraised included goal setting, social opportunities, and progress monitoring. Benefits of the intervention, including increased levels of exercise; improved diet and physical health; increased psychological wellbeing (e.g. confidence, self-esteem); and improved social relationships, were identified by participants, independent of actual weight loss. |
| Predictors of Attendance in Health and Wellness Treatment Groups for People with Serious Mental Illness | Phalen et al | 2020 | US | Cluster search on study ID | Psychiatric diagnosis of schizophrenia spectrum disorder, bipolar disorder, major depression with psychotic features, and/or post-traumatic stress disorder, and a cooccurring chronic medical condition. | Secondary analysis of RCT data | We found lower attendance among people who were younger, people with more medical conditions, and people with more emergency room visits in the six months prior to the beginning of treatment. Younger age was a particularly strong predictor of low attendance and was the only variable significantly associated with attending zero treatment sessions. |

| | | | | | | | |
|---|---|---|---|---|---|---|---|
| The Scandinavian Solutions for Wellness study - a two-arm observational study on the effectiveness of lifestyle intervention on subjective well-being and weight among persons with psychiatric disorders | Porsdal et al | 2010 | Denmark, Norway and Sweden | Medline | Psychiatric diagnosis and being on psychotropic medication, defined as antipsychotics, antidepressants or mood stabilizers | Observational study | 314 patients enrolled in the SfW group, 59 in the control group. 54% of the patients had schizophrenia, 67% received atypical antipsychotics, 56% were female. They averaged 41 ± 12.06 years and had a BMI of 31.4 ± 6.35. There were significant differences at baseline between groups for weight, SWN total score and other factors. Stepwise logistic models controlling for baseline covariates yielded an adjusted non-significant association between SfW program participation and response in subjective well-being (SWN increase). However, statistically significant associations were found between program participation and weight-response (weight loss or gain < 1 kg) OR = 2; 95% CI [1.1; 3.7] and between program participation and WC-response (WC decrease or increase < 2 cm) OR = 5; 95% CI [2.4; 10.3]), at 3 months after baseline. |
| Incentives and barriers to lifestyle interventions for people with severe mental illness: a narrative synthesis of quantitative, qualitative and mixed methods studies | Roberts & Jois | 2011 | UK | Medline | Diagnosis of severe mental illness, psychosis, schizophrenia, schizophreniform disorder, schizoaffective disorder or bipolar disorder | Narrative synthesis | No studies were identified that specifically explored the incentives and barriers to participation in lifestyle intervention for this population. Existing literature report some possible incentives and barriers including: illness symptoms, treatment effects, lack of support and negative staff attitudes as possible barriers; and symptom reduction, peer and staff support, knowledge, personal attributes and participation of staff as possible incentives. |

| | | | | | | | |
|---|---|---|---|---|---|---|---|
| An ethnographic study of the incentives and barriers to lifestyle interventions for people with severe mental illness | Roberts & Jois | 2013 | UK | Medline | Mental health service users | Ethnographic qualitative study | Participant observation highlighted environment, facilitator style, group ownership, group cohesion, information and learning, incentives and barriers as important. Participant interviews identified weight management, social networking, information and communication, role of healthcare professionals and perceived benefits as key themes. |
| Health Improving Measures for Patients Suffering from Psychosis Development of treatment methods and an intervention program | Sameby et al | 2008 | Sweden | CINAHL | Psychosis | Experimental action research | Patients identified improved diet, weight loss, and increased physical activity as prioritized goals. 14 patients completed the entire program and 8 of these (67%) had a lower BMI. A further reduction was seen in 5 persons at the 18 month follow-up but 6 others (43%) had increased BMI. In response to the open-ended evaluation questions, patients reported increased social contact and improved health awareness. |
| Disordered Eating among People with Schizophrenia Spectrum Disorders: A Systematic Review | Sankaranarayanan et al | 2021 | Australia | Via project team | Patients with a diagnosis of schizophrenia, schizoaffective disorder, schizophreniform disorders and first episode schizophrenia or includes Schizophrenia spectrum disorders | Systematic review | The ma-jority of studies (17) rated fair on qualitative analysis and included more men, and participants in their 30s and 40s, on antipsychotics. The commonest limitations include lack of sample size or power calculations, poor sample description, not using valid tools, or not adjusting for con-founders. The reported rates were 4.4% to 45% for binge eating, 16.1% to 64%, for food craving, 27% to 60.6% for food addiction, and 4% to 30% for night eating. Positive associations were re-ported for binge eating with antipsychotic use and female gender, between food craving and weight gain, between food addiction and increased dietary intake, and between disordered eating and female gender, mood and psychotic symptoms. Reported rates for disordered eating among people with SSD are higher than those in the general population. |

| | | | | | | | |
|---|---|---|---|---|---|---|---|
| Perspectives on Diet and Physical Activity among Urban African Americans with Serious Mental Illness | Sayer et al | 2019 | US | Citation searching from Lee et al, 2022 | Serious mental illness - mental health service user | Qualitative (incorporating descriptive statistics) | This paper discussed findings on the perspectives of urban African Americans with SMI related to diet, physical activity, and obesity. By and large, participants viewed obesity as the cause of many health problems, understood that diet and physical activity were important components of their health, and believed that health and weight of African Americans with SMI are shaped by many factors.<br><br>Structural barriers were discussed as hurdles to healthy eating and physical activity. This group of African Americans with SMI articulated many of the same structural barriers common to persons in African American and low-income communities. These include a lack of safe places to exercise, limited income that makes healthy food and gym membership unaffordable, and poor access to supermarkets and healthy food environments (Baker et al., 2006; Gordon-Larsen et al., 2006). Study findings emphasize the need for weight loss interventions for African Americans with SMI to attend to structural challenges and find ways to surmount them. |
| Learning what matters for patients: qualitative evaluation of a health promotion program for those with serious mental illness | Shiner et al | 2008 | US | Citation searching from Lee et al, 2022 | Serious mental illness | Appreciative enquiry | Among successful participants, three themes emerged, highlighting the importance of: (i) individualized interventions promoting engagement in the program; (ii) relationships with health-promotion program employees and (iii) self-confidence resulting from program participation. Health-promotion programs that target these areas may have better success in achieving health benefits for persons with SMI. |

| | | | | | | | |
|---|---|---|---|---|---|---|---|
| Early Intervention in Psychosis: Effectiveness and Implementation of a Combined Exercise and Health Behavior Intervention Within Routine Care | Smith et al | 2020 | UK | Medline | First episode psychosis | Non-randomised clinical trial | Mean baseline data suggests participants were at an increased health risk on entry to the program, with elevated values in mean body mass index (BMI; 70% overweight/obese), waist circumference, resting heart rate, and triglycerides. Fifty percent reported smoking daily, 64% ate < 5 fruits/vegetables per day, and 52% of participants were prescribed highly obesogenic antipsychotic medications (i.e., Olanzapine). At 12 weeks and 12 months, no changes were observed in mean BMI, waist circumference or any other clinical variable ($p > 0.05$). At 12 months, participants reported a positive impact on health behaviors including improved diet, increased physical activity levels, and cessation of substance use (n = 2), alcohol use (n = 2), and smoking (n = 4). Focus groups captured participant experiences, engagement with and satisfaction with the program, including challenges/barriers to program adherence. |
| Strategies and Lessons Learned for Supporting and Supervising Peer Specialists | Stefancic et al | 2021 | US | Cluster search on study ID | Serious mental illness - Peer specialists | Qualitative: Reflection on delievry of an RCT | Strategies included access to multiple supervisors, formal and informal support, acknowledgment of lack of role clarity, ongoing role negotiation, a collaborative approach to troubleshooting challenges, explicit emphasis on peer specialists' value, and linking peer specialists to opportunities for mutual support and professional development. |

| | | | | | | | |
|---|---|---|---|---|---|---|---|
| Feasibility and Acceptability of a Lifestyle Intervention for Individuals with Bipolar Disorder | Sylvia, Janos et al | 2019 | US | Cluster search on study ID | Primary diagnosis of bipolar disorder (Type I or II) | RCT | There was no statistically significant difference in drop-out rates between the groups (26.3% in NEW Tx, 31.6% in the control condition). In the NEW Tx condition, participants attended a mean of 66.7% of sessions and reported moderate to high satisfaction. There were no study-related adverse events. We also found that expectations, but not perceived credibility (or believability), of NEW Tx (as measured by the Credibility/ Expectancy Questionnaire) at baseline predicted treatment satisfaction (as measured by the Care Satisfaction Questionnaire) post-treatment. Manic symptoms at baseline predicted treatment satisfaction, and marital status predicted one's expectations of lifestyle interventions. |
| Pilot Study of a Lifestyle Intervention for Bipolar Disorder: Nutrition Exercise Wellness Treatment (NEW Tx) | Sylvia, Pegg et al | 2019 | US | Citation searching from Lee et al, 2022 | Primary diagnosis of BD (Type I or II) | Pilot study | The NEW Tx group reported increased weekly exercise duration and overall functioning, and decreased depression and illness severity over the study duration. However, only improvements in functioning were significantly greater in the NEW Tx group than in the control group. There were no group differences in weight loss or mood symptoms over the study duration. |
| Correlates of Attendance in a Peer-Led Healthy Lifestyle Intervention for People with Serious Mental Illness Living in Supportive Housing | Tuda et al | 2021 | USA | Cluster search on study ID | Serious mental illness diagnosis | Secondary analysis of RCT | Results indicated that females, those with at least a high school education, and a diagnosis of schizophrenia were more likely to attend. In contrast, the odds of attending at least one session were significantly lower for those who reported any drug use and for those who rated their health as good or excellent. |

| | | | | | | | |
|---|---|---|---|---|---|---|---|
| The experience of weight gain as a result of taking second-generation antipsychotic medications: the mental health consumer perspective | Usher et al | 2013 | Australia | Cluster search on author | Schizophrenia | Qualitative | Thematic analysis resulted in three themes: Grappling with the weight; Living with the consequences of being overweight; and Experiencing negative emotions about the weight gain. The findings indicate that consumers struggle to manage the insatiable appetite and the related weight gain associated with second-generation antipsychotic medication, as well as the numerous associated physical and emotional issues. Adherence with prescribed second-generation antipsychotic medication was also affected and a number of the participants indicated they had ceased or considered ceasing their medication because of the weight gain associated with the drugs. |
| A Pilot Evaluation of the In SHAPE Individualized Health Promotion Intervention for Adults with Mental Illness | Vancitters et al | 2010 | US | Cluster search on author | primary DSM-IV Axis I or Axis II diagnosis | Pilot study | Over a 9-month period, participation was associated with increased exercise, vigorous activity, and leisurely walking ($P<.01$), and a trend toward improved readiness to reduce caloric intake ($P = .053$). Participants demonstrated a reduction in waist circumference ($P<.05$), but no change in BMI. Satisfaction with fitness ($P<.001$) and mental health functioning ($P<.05$) improved, and severity of negative symptoms decreased ($P<.01$). This study demonstrated the feasibility and potential effectiveness of the In SHAPE program, which consisted of exercise and dietary modification. |

| | | | | | | | |
|---|---|---|---|---|---|---|---|
| Qualitative descriptive study exploring schizophrenia and the everyday effect of medication-induced weight gain | Vandyk & Baker | 2012 | USA | CINAHL | diagnosis of schizophrenia or schizoaffective disorder | Qualitative | Data were analysed according to the method of constant comparison and three central themes emerged: a life altering diagnosis, weight management as complex, and today's experiences shape tomorrow's outcomes. Weight management was seen as difficult yet important to the participants. The findings of this study provide insight into the views and opinions of the participants regarding weight and lifestyle and may be used to support the design of tailored heath initiatives for persons with mental illness. |
| Perceptions of Strategies for Successful Weight Loss in Persons with Serious Mental Illness Participating in a Behavioral Weight Loss Intervention: A Qualitative Study | Vazin et al | 2016 | USA | Citation searching from Lee et al, 2022 | Serious mental illness | Qualitative | Participants perceived tailored exercise sessions, social support, and dietary strategies taught in ACHIEVE – such as reducing portion sizes and avoiding sugar-sweetened beverages – as useful weight loss strategies. Health benefits, improved physical appearance, self-efficacy, and enhanced ability to perform activities of daily living were commonly cited benefits of intervention participation and weight loss. Some participants reported challenges with giving up snack food and reducing portion sizes, and barriers to exercise related to medical conditions. |

| | | | | | | | |
|---|---|---|---|---|---|---|---|
| Health promotion in individuals with mental disorders: a cluster preference randomized controlled trial | Verhaeghe et al | 2013 | Belgium | Citation from Mucheru | Mental disorders, including schizophrenia, bipolar disorders, depression, and anxiety disorders | RCT | A significant difference was found between the intervention group and the control group regarding body weight (−0.35 vs. +0.22 kg; p=0.04), Body Mass Index (−0.12 vs. +0.08 kg/m2; p=0.04), waist circumference (−0.29 vs. + 0.55 cm; p<0.01), and fat mass (−0.99 vs. −0.12%; p<0.01). The decrease in these outcomes in the intervention group disappeared during the follow up period, except for fat mass. Within the intervention group, a larger decrease in the primary outcomes was found in the participants who completed the intervention. No significant differences between the two groups in changes in the secondary outcomes were found, except for the pedometer-determined steps/day. In the intervention group, the mean number of daily steps increased, while it decreased in the control group. |
| Health promotion in mental health care: perceptions from patients and mental health nurses | Verhaege et al | 2013 | Belgium | Citation from Mucheru | Mental disorders: schizophrenia, schizoaffective disorders, bipolar and depressive disorders | Qualitative | Although physical and mental health benefits of physical activity and healthy eating were identified, several barriers to integrate healthy lifestyles into the daily life of patients were reported. Important barriers identified by the patients consisted of lack of energy and motivation as a result of the MD, side effects of psychotropic drug use, and hospitalisation. Lack of time and personal views and attitudes towards health promotion were reported by the mental health nurses as important elements influencing the way in which they integrate health promotion in the care provided. Support from the mental health nurse was considered important by the patients in changing their unhealthy lifestyle behaviour. |

| | | | | | | | |
|---|---|---|---|---|---|---|---|
| The psychological journey of weight gain in psychosis | Waite et al | 2022 | England | Personal Contacts | primary diagnosis of non-affective psychosis (e.g., schizophrenia, schizoaffective disorder, schizophreniform disorder, delusional disorder, brief psychotic disorder and psychotic disorder not otherwise specified) | Qualitative | Patients described that initially the extent and speed of weight gain was overshadowed by psychotic experiences and their treatment. This led to a shocking realisation of weight gain. The psychological impact of weight gain, most strikingly on the self-concept, was profound. Loss of self-worth and changed appearance amplified a sense of vulnerability. There were further consequences on mood, activity and psychotic experiences, such as voices commenting on appearance, that were additional obstacles in the challenging process of weight loss. Sedative effects of medication also contributed. Unsuccessful weight loss left little hope and few preferences for interventions. Early information about common weight gain trajectories and working with experts-by-experience were valued. Rebuilding self-confidence, efficacy and worth may be a necessary first step. |
| Keeping the body in mind: A qualitative analysis of the experiences of people experiencing first-episode psychosis participating in a lifestyle intervention programme | Watkins et al | 2020 | Australia | CINAHL | DSM diagnoses including schizophreniform, bipolar affect disorder, depression with psychotic features, and schizophrenia | Qualitative | Eleven participants were interviewed (seven males), aged between 18 and 25 years. Thematic analysis revealed four main themes: the role of physical health in mental health recovery; the importance of staff interactions; the value of peer interaction; and graduation to a sustainable healthy lifestyle. Study participants reported that they valued the programme for both their |

| | | | | | | | |
|---|---|---|---|---|---|---|---|
| | | | | | | | physical health<br>andmental health recovery. |
| 'Walk This Way' – a pilot of a health coaching intervention to reduce sedentary behaviour and increase low intensity exercise in people with serious mental illness: study protocol for a randomised controlled trial | Williams et al | 2016 | UK | Cluster search on author | Diagnosis of any SMI (schizophrenia, psychosis, bipolar disorder and major depression) | Pilot RCT - Protocol | N/A |
| Psychosocial processes influencing weight management among persons newly prescribed atypical antipsychotic medications | Xiao et al | 2012 | Canada | CINAHL | Newly prescribed atypical antipsychotic medications | Qualitative (incorporating descriptive statistics) | Contextual factors influencing weight management were: accessibility to resources, unstructured lifestyle, and others' perception of weight. Conditions influencing weight management were: rapid weight gain, insatiable hunger and lack of motivation boosters. Participants' early responses to weight gain included discontinuing medications, choosing lower-calorie foods, using walking in daily activities as exercise, accepting weight gain and trying to manage weight but giving up. The consequences revealed from data analysis were contemplating weight management and not trying, as the barriers to weight management exceeded the facilitators. |

| | | | | | | | |
|---|---|---|---|---|---|---|---|
| Improving Lifestyle Interventions for People With Serious Mental Illnesses: Qualitative Results From the STRIDE Study | Yarborough et al | 2016 | US | Citation searching from Lee et al, 2022 | Serious mental illness | Qualitative (subset of RCT data) | Barriers to behavior change were similar to those described for the general population, including lack of support from significant others, the lure of unhealthy foods, and poor weather impeding exercise. Additional challenges included the effects of psychiatric symptoms, or consequences of symptoms (i.e., social isolation), on ability to make and sustain lifestyle changes. We found a strong preference for ongoing, group-based support to foster a sense of accountability which motivated and helped to sustain behavior changes. |
| Patient perspectives on how living with a mental illness affects making and maintaining healthy lifestyle changes | Yarborough et al | 2019 | US | Cluster search on author | Schizophrenia spectrum disorders, bipolar disorder/affective psychoses, major depressive disorder, or anxiety disorders | Qualitative | Three themes described patients' (n=163) perspectives on barriers to making healthy lifestyle changes: 1) Thinking about making lifestyle changes is overwhelming for individuals already managing the burdens of mental illnesses; 2) Depression makes it difficult to care about a healthy future; and 3) When mental illness symptoms are not adequately treated unhealthy behaviors that provide relief are unlikely to be discontinued. Participants also made suggestions for improving health care delivery to facilitate positive behavior change. |

| | | | | | | | |
|---|---|---|---|---|---|---|---|
| Improving Weight in People with Serious Mental Illness: The Effectiveness of Computerized Services with Peer Coaches | Young et al | 2017 | US | Citation searching from Lee et al, 2022 | diagnosis of schizophrenia, schizoaffective disorder, bipolar disorder, major depressive disorder with psychosis, or posttraumatic stress disorder | RCT | At 6 months, in obese patients (n=200), there was a significant condition by visit effect (F = 4.02, p = 0.02). TheWebMOVE group had an average estimated BMI change from baseline to 6 months of 34.9 ± 0.4 to 34.1 ± 0.4. This corresponds to 2.8 kg (6.2 lbs) weight loss (t = 3.2, p = 0.001). No significant change in BMI was seen with either in-person services (t=0.10, p = 0.92), or usual care (t = −0.25, p = 0.80). The average percentage of modules completed in the WebMOVE group was 49% and in the in-person group was 41% (t=1.4, p = 0.17). When non-obese patients were included in the analyses, there was a trend towards a condition by visit effect (F=2.8, p = 0.06). WebMOVE was well received, while the acceptability of in-person services was mixed. |

#### Supplementary File 3: RAMESES Checklist for Realist Reviews

Completed 29th October 2024

**1** - In the title, identify the document as a realist synthesis or review

*See title*

ABSTRACT

**2** - While acknowledging publication requirements and house style, abstracts should ideally contain brief details of: the study's background, review question or objectives; search strategy; methods of selection, appraisal, analysis and synthesis of sources; main results; and implications for practice.

*See Abstract*

INTRODUCTION

**3** - Rationale for review - Explain why the review is needed and what it is likely to contribute to existing understanding of the topic area.

*See Introduction Pg 5-6*

**4** - Objectives and focus of review - State the objective(s) of the review and/or the review question(s). Define and provide a rationale for the focus of the review.

*See Introduction, Pg 5-6*

METHODS

**5** - Changes in the review process - Any changes made to the review process that was initially planned should be briefly described and justified.

*See Pg 6-, Stage 1: Developing the initial programme theory*

*See Pg 7, Stage 2: Searching for evidence*

*See Pg 8, Stage 3: Selection, appraisal and data extraction*

*See Pg 10 Stage 5:Data synthesis, CMOC development and programme theory refinement*

**6** - Rationale for using realist synthesis - Explain why realist synthesis was considered the most appropriate method to use.

*See Pg 5, Introduction,*

**7** - Scoping the literature - Describe and justify the initial process of exploratory scoping of the literature.

*See Pg 6, Developing the initial programme theory*

**8** - Searching processes - While considering specific requirements of the journal or other publication outlet, state and provide a rationale for how the iterative searching was done. Provide details on all the sources accessed for information in the review. Where searching in electronic databases has taken place, the details should include, for example, name of database, search terms, dates of coverage and date last searched. If individuals familiar with the relevant literature and/or topic area were contacted, indicate how they were identified and selected.

*See Pg 2, Stage 2: Searching for evidence*

*See Supplemental file 1*

**9** - Selection and appraisal of documents - Explain how judgements were made about including and excluding data from documents, and justify these.

*See pg 8 &9, Stage 3: Screening and inclusion*

**10** - Data extraction - Describe and explain which data or information were extracted from the included documents and justify this selection.

*See Pg 10 – Stage 4: Data Extraction and Analysis*

**11** - Analysis and synthesis processes - Describe the analysis and synthesis processes in detail. This section should include information on the constructs analyzed and describe the analytic process.

*See pg 10– Stage 4: Data extraction, analysis and Stage 5: Data synthesis, CMOC development and programme theory refinement*

RESULTS

**12** - Document flow diagram - Provide details on the number of documents assessed for eligibility and included in the review with reasons for exclusion at each stage as well as an indication of their source of origin (for example, from searching databases, reference lists and so on). You may consider using the example templates (which are likely to need modification to suit the data) that are provided.

*See PRISMA diagram, Figure 1*

**13** - Document characteristics - Provide information on the characteristics of the documents included in the review.

*See Supplementary file 2*

**14** - Main findings - Present the key findings with a specific focus on theory building and testing.

*See Results section*

*See Supplementary File 3*

DISCUSSION

**15** - Summary of findings - Summarize the main findings, taking into account the review's objective(s), research question(s), focus and intended audience(s).

*See Pg 25, Discussion, paragraph 1: Key Findings*

**16** - Strengths, limitations and future research directions - Discuss both the strengths of the review and its limitations. These should include (but need not be restricted to) (a) consideration of all the steps in the review process and (b) comment on the overall strength of evidence supporting the explanatory insights which emerged.

The limitations identified may point to areas where further work is needed.

*See pg 31, Limitations & Strengths*

**17** - Comparison with existing literature - Where applicable, compare and contrast the review's findings with the existing literature (for example, other reviews) on the same topic.

*See Pg 26 - 30, Comparable Literature to Support the Programme Theory and CMOCs*

**18** - Conclusion and recommendations - List the main implications of the findings and place these in the context of other relevant literature. If appropriate, offer recommendations for policy and practice.

*See Pg 32, Conclusion*

**19** – Funding - Provide details of funding source (if any) for the review, the role played by the funder (if any) and any conflicts of interests of the authors.

*Funding disclaimer noted in the online portal*

*See page 1 for conflicts of interest from the authors*

#### RESOLVE Review Supplementary File 4: MEDIATE CMOC data appendix

Each CMOC developed during the review is presented below with the underpinning exemplar evidence (extracted text from papers complete with existing references if appropriate). A reference list for the exemplar evidence can be found at the end of this document

Bold portions of text reflect researcher perceived level of importance at time of initial CMOC composition

##### CMOC 1 ('Building therapeutic relationships with professionals')

**When individuals with SMI and anti-psychotic weight gain develop therapeutic relationships with non-judgemental, honest professionals who objectively explain the importance of interventions in lay terms (C), individuals are more likely to follow the advice and engage with the interventions (O), because they feel respected by professionals (M) and trust that professionals apply expert knowledge to their needs (M)**

CMOC based on data extracted from 10 documents.

| Document (author, date) | Extracted data |
|---|---|
| (Bochicchio et al., 2021) | Participants [SUs] emphasized that a **long history of working with a particular NPS** [non-peer support] **fostered a sense of trust and familiarity.** *"It's been three years…I get an honest response from her…I know she's not lying to me."*<br>Overall, non-peer providers were **appreciated for their expertise in the domain of health, their ability to translate health information into understandable terms, and the directness of their feedback**, with some exceptions.<br>*…" they know what they're talking about…they're doctors, and they went to school for things. And if they say I need to lose a couple of pounds, then they're right…the doctor is more focused on…the objective sides…blood pressure, vital signs, blood work."*<br><br>One participant explained,<br>*"That's the biggest thing…**He did put it in things I could understand**, but because he worded it certain ways, **I was able to relate more to my own problems.** I was able to communicate my own problems better because it was in **simple, plain English**."*<br>Another participant articulated the importance of their NPS's expertise in conveying health data, *"she showed me the levels of diabetes in my blood work…she took her time to explain what each level meant…they have different signs and signatures on the graph…but **she helped me understand."*** |
| (Nover, 2013) | **Several respondents described not adhering to their usual treatment regimen because they did not feel like their opinions were valued. After participating in the CPCI program, participants stated that they felt less judged, more familiar with the** |

| | |
|---|---|
| | **providers at the clinic and more knowledgeable about healthy living**—all factors that can contribute to a more positive health care experience |
| (Park et al., 2017) | An essential component of mental health care is the **development of therapeutic relationships** (Akerjordet & Severinsson, 2004). This relationship is fundamental to the role of mental health professionals and includes consideration of **all aspects of the person**. The partnership developed within **the therapeutic relationship facilitates the achievement of individual health goals within a supportive and authentic relationship (**Akerjordet &Severinsson, 2004)[64] |
| (Verhaeghe et al., 2013) | Support from the mental health nurse was considered important by the patients in changing their unhealthy lifestyle behaviour. Support and discussing lifestyle issues was considered important and desirable by a majority of the participating **patients because it made them feel respected as an individual.** They stated that, by asking them about lifestyle issues, mental health nurses showed interest. Moreover, some patients described that they indeed would follow the advice of the mental health nurse: **When she [nurse] gives me advice how to cook using healthy ingredients or to go to the fitness, I would follow this advice. The fact that she mentions it means for me that here is a problem. In this sense, I would follow her advice** |

#### CMOC 2 ('The scope of practitioners')

**When practitioners working with individuals with SMI and weight gain from anti-psychotic medications have specialized knowledge of weight management challenges associated with SMI medications (C), proactive assessment and weight management are more likely to be incorporated into individuals' treatment plans (O), due to practitioners' awareness of critical mental and physical health indicators (M).**

CMOC based on data extracted from 16 documents.

| Document (author, date) | Extracted data |
|---|---|
| (Chalfoun, 2016) | …one of the main goals of this review is to **encourage health professionals in testing and improving cardiorespiratory fitness levels as well as waist circumference in individuals with schizophrenia**. Indeed, one of the main examples of a significant clinical implication that can be drawn from this review relates to the improvement of cardiorespiratory fitness and waist circumference, which was attained in most studies |
| (Forsberg et al., 2008) | The present study also emphasizes the importance of a **comprehensive view in the care of persons with psychiatric disabilities**. Professionals in the psychiatric field should in their daily meetings with persons with psychiatric disabilities aim to increase the understanding of the importance of a healthy lifestyle and how it can reduce the risk of untimely illness and death. The professionals also have to support the mentally ill when making decisions both in the short and the long term and motivate the promotion of health and a life-long change in lifestyle |
| (Kreyenbuhl et al., 2019) | To give patients more realistic expectations about medication side effects, **prescribers should endeavor to have well-informed, patient-centered, individualized, and mutually respectful discussions proactively about medication side effects and the many** |

| | |
|---|---|
| | **uncertainties associated with them.** These discussions should include **explicit evaluations** of veterans' willingness to accept certain side effects like weight gain, assessment of other risk and protective factors regarding fitness and weight, and prevention strategies. They should incorporate emotional support for dealing with the downside of the trade-off and limited options |
| (Lee et al., 2023) | …In the case of obesity, this has previously been attributed to lack of training and confidence around management.20 **There may also be a lack of proactiveness from healthcare professionals, with the condition going unrecognised in patients' clinical notes, clinicians underestimating the severity of patients' excess weight**, clinicians lacking awareness of treatment options and having a negative attitude towards their efficacy, and the perception among clinicians that obesity arises from lifestyle choices with weight management being primarily the responsibility of the patient.11,21. |

#### CMOC 3 ('Coordinated/integrated care approaches')

**When those involved in caring for individuals with SMI and anti-psychotic induced weight gain offer consistent contact and work together with individuals to assess and monitor their care needs (C), co-developed goals are more likely to be met (O), due to greater shared responsibility between individuals and key services (M)**

CMOC based on data extracted from 3 documents.

| Document (author, date) | Extracted data |
|---|---|
| (Bochicchio et al., 2021) | Individuals with severe mental illness (SMI) face significant health disparities and multiple barriers to engaging in health behaviour change. To reduce these health disparities, it is necessary to enhance the support individuals with SMI receive through the **collaboration of different healthcare providers.**<br>**…Integrated care models** seeking to maximize collaboration between physical and mental health providers (e.g., medical doctors, social workers, case managers, paraprofessional staff) have gained traction to counteract reliance on separate specialists providing treatment in silos (Rodgers et al., 2016). Such approaches are associated with improved care quality and physical health outcomes (e.g. improvements in cholesterol), and lower overall costs (Walker & Druss, 2018; Scharf et al., 2016). |
| (Goodrich et al., 2013) | Current CCMs (collaborative care models) are an iteration of the Chronic Care Model that acknowledges mental disorders also require a long-term and systematic approach to foster access and continuity of care to achieve optimal management. Moreover, mental health CCMs **emphasize collaboration among a team of mental health providers and PCPs within a practice to effect these changes, including coordination of care with specialists and community resources outside of primary care**…. **Care management** is a key operational component of CCM health care system redesign and represents a significant change from traditional physician-centered, primary care practice [12, 24, 86]. In the CCM practice environment, PCPs (primary care providers) are part of a team and are responsible for the screening and diagnosis of mental health conditions, prescribing |

| | |
|---|---|
| | appropriate medications, and referring complex cases to specialty mental health care as needed. **PCPs delegate and supervise many treatment tasks, which are coordinated by the care manager, to other members of the care team. Physicians are indirectly supported by mental health specialists, such as psychiatrists, who provide decision support for complex cases, as well as treatment recommendations** [7, 32]… Several systematic, meta-analytic reviews were published over the last year that provided robust support for CCMs as an **evidenced-based strategy for the management of mental health conditions in primary care settings** |

#### CMOC 4 ('Important assessment considerations: addressing negative body image')

**When individuals with SMI and weight gain from anti-psychotic medications have negative body image due to weight stigma, skilled professionals can help 'reframe' their situation (e.g. using functionality-based techniques) (C), potentially decreasing weight stigma and increasing positive body image (O) due to individuals' greater appreciation of themselves as holistic human beings (M).**

CMOC based on data extracted from 3 documents.

| Document (author, date) | Extracted data |
|---|---|
| (Waite et al., 2022) | Interventions may also need to address issues concerning current **societal stigma** regarding both psychotic experiences and excess weight, with the potential to draw on the **emerging body positivity movement** ideas concerning more compassionate approaches. |
| (Mahfoud et al., 2023) | …**interventions aimed at improving functionality appreciation** could be regarded beneficial therapeutic targets in the treatment of psychosis. Previous studies demonstrated that functionality-based techniques (e.g., body functionality structured writing exercises) are **effective in reducing beliefs that the body is valued positively only through physical appearance, and in promoting a more holistic perception of the body** [34, 60]. |

#### CMOC 5 ('Important assessment considerations: addressing existing disordered eating')

**When SUs with SMI and weight gain from anti-psychotic medications have existing disordered eating behaviours, early assessment by skilled professionals (C), may result in a more effective obesity management plan (O), due to individual and practitioner awareness (M) of how disordered eating behaviours can negatively impact an individual's mental and physical health**

CMOC based on data extracted from 4 documents.

| Document (author, date) | Extracted data |
|---|---|
| (Alliende & Mittal, 2023) | Any plan to prevent weight gain, promote weight loss, or install new health behaviours more generally in people with schizophrenia needs to consider that it will be dealing with a population where eating and weight-related beliefs and behaviours are more likely to be altered. **Screening patients with a schizophrenia diagnosis for potential eating disorders should be incorporated as part of designing a treatment plan for weight management or metabolic health** |
| (Barre et al., 2011) | Clinical practitioners should inquire about eating habits and empower individuals to eat a healthier diet. **Clinicians could also ask about eating in response to emotions and the impact of antipsychotic medication side effects on eating behaviours**. Through collaborative efforts and refocused attention on nutrition in the mental and medical health sectors, the diet and health of consumers may be improved….For those ready to make changes, **nutrition education should begin with defining healthy diets, evaluating the quality of individual diets, and exploring personal health risks from unhealthy eating**. Moderation in eating of all foods and establishment of lifelong healthy eating habits should be emphasized versus the notion of dieting and elimination of foods. Education efforts should be individualized and include planning and preparation of meals within cost and cooking limitations. Opportunities to taste and prepare healthy foods, particularly convenient healthy foods, could be incorporated. Registered dietitians could aid in nutrition education |
| (Sankaranarayanan et al., 2021) | …when creating weight management, weight gain prevention, or metabolic health plans with patients**, it is important to take mental health into consideration in the form of comorbid eating disorders. Notably, people with schizophrenia have higher rates of binge eating disorder** than the general population (approximately 10%, compared to 0.7–4.3% in community sample) (Kouidrat, Amad, Lalau, & Loas, 2014). …abnormal eating behaviours that do not meet criteria for an eating disorder – such as food craving, food addiction, or night eating were also found to be highly prevalent in people with schizophrenia – 2.5 to 4 times higher than in the general population…. **As the rates for disordered eating are higher among people with schizophrenia spectrum disorder [SSD], it is important to train clinicians to look for, assess and manage eating disorders at various stages of the illness**, particularly modelled along the lines of those for substance-use disorders [56]. |

#### CMOC 6 ('Behaviour change strategies to increase readiness for participation')

**When individuals with SMI and weight gain from anti-psychotic medications receive early behaviour change support from skilled professionals (C), they are more apt to consider participation in weight management options (O), because they have increased perceptions of self-efficacy (SE) (M).**

CMOC based on data extracted from 15 documents.

| Document (author, date) | Extracted data |
|---|---|
| (Bennett et al., 2018) | Taken together, prior research and findings from this study indicate that **the relationship between SE [self-efficacy] and participation is robust and should be considered when designing and implementing weight management interventions for individuals with SMI.** Including a **short intervention bolstering SE** (i.e., **motivational interviewing** focused on identifying prior displays of SE and successful change) before the weight management intervention may be beneficial (Armstrong et al., 2011). |
| (Casagrande et al., 2010) | The theoretical base of the ACHIEVE Trial fits well within the psychiatric rehabilitation framework which emphasizes tenets of intrinsic skills building and environmental support [37,38]. **Motivational interviewing provides an important framework for helping participants problem solve and set goals for weight loss.** The ACHIEVE intervention operationalizes these models by providing frequent and extended contacts, opportunities for group interactions and social support, goal setting and self-negotiation, problem solving, and examples of new behavioural options |

#### CMOC 7 ('Early-to-maintenance phases of weight management')

**When individuals with SMI and weight gain from anti-psychotic medications are offered flexible program options (e.g., individual, group sessions) using an individualised 'small steps' approach by skilled practitioners (C), individuals are more apt to begin co-developing relevant, manageable behaviour change goals (O), because these approaches are easier to integrate into daily routines, building on individuals' sense of achievement (M).**

CMOC based on data extracted from 29 documents.

| Document (author, date) | Extracted data |
|---|---|
| (Bartels et al., 2015) | Results from these three studies confirm the effectiveness of lifestyle interventions for overweight and obese persons with severe mental illness in achieving clinically significant reduction in cardiovascular risk either **through group or individual coaching, and for different degrees of intensity of dietary and exercise programming** |
| (Pedley et al., 2018) | **Participants described how setting goals motivated positive changes to lifestyle and provided a framework by which to gradually 'step up' adjustments in a manageable way:** |

| | |
|---|---|
| | *"… Once you reach the three-month goal, it gives you this spur to go for the six months…"* **(Pt.8)** |
| (Shiner et al., 2008) | Most lifestyle interventions are relatively short and intense programmes that usually have **at least three key components**: **exercise, diet and behavioural therapy [3, 4**]. Although these interventions are effective in the short term, the benefits are seldom sustained in the long term. The literature shows that while interventions aimed at making small but sustainable changes in lifestyle behaviour – **the 'small steps approach'**– not only lead to small weight changes in the short term, **they prevent weight regain and result in more structural weight loss in the long term because such small changes can be sustained** [5–7]. The small-steps approach is of interest to clinical care, insofar as it might be **easier to implement in the daily care of patients than other interventions**. It may also be suitable for any patient and not just those who are highly motivated and stable |
| (Aschbrenner et al., 2016) | **The In SHAPE health coach works with each participant to develop personalized lifestyle and fitness evaluations**, and meets weekly with participants for 1-hour sessions at a local gym (e.g., YMCA). During the gym sessions, the health coach provides supported fitness coaching and **individualized attention to the participant's nutrition goals and objectives**. The coaches also provide participants with support for managing mental health symptoms that interfere with exercise and healthy eating….At the start of the program, the health mentors meet with participants to conduct comprehensive lifestyle and fitness evaluations and develop personalized fitness plans for each participant with **shared goal setting. Helping participants translate lessons learned during individual coaching sessions at a fitness facility to their home and social environments might be critical for long-term weight loss success** |
| (Bergqvist et al., 2013) | The participants in this study [practitioners] described how to begin the **exploration of daily routines** in order to identify health factors such as diet, exercise, smoking habits, and alcohol consumption, with **the purpose being to collaborate with the patient and see what could or needed to be changed….** Participants in this study stated that **changes must be adapted to what is important for the individual**. **Information must continue to be individualized and concrete, and practical examples** should be given when the individuals need them |

#### CMOC 8 ('Maintenance-to transition phases of weight management')

**When individuals with SMI and weight gain from anti-psychotic medications have ongoing, individualized support from empathic, trained practitioners and/or peer mentors to better manage their specific life challenges and opportunities (C), individuals are more likely to transition to autonomous goal-setting and self-monitoring (O), because they feel empowered by their increased perceptions of control over lifestyle management (M).**

CMOC based on data extracted from 12 documents.

| Document (author, date) | Extracted data |
|---|---|
| (Bergqvist et al., 2013) | The results from this study also highlight the importance of **identifying opportunities and obstacles** for the individuals… Communication and empathy are prerequisites for a good interpersonal relationship (Travelbee,1971). **Empathy** is characterized by |

| | |
|---|---|
| | the ability to share another individual's experiences and thereby understand his or her thoughts and feelings. This requires self-awareness and an understanding of the individual's actions and how to interpret them. These are characteristics that participants in this study perceive that they use in their work |
| (Bochicchio et al., 2021) | In a recent systematic literature review examining health outcomes among persons with SMI receiving services in behavioural health homes, the inclusion of **peer support and/ or training in self-management skills** were associated with the greatest reduction in cardiometabolic risk factors |
| (Smith et al., 2020) | Maintenance of health behaviours and long-term sustainability were identified as key indicators of success. Exercise program design was grounded using aspects of self-determination theory (43, 44), including **key strategies structured to foster PA [physical activity] self-efficacy and improve perceived competence by offering a range of mastery experiences with education about monitoring exercise intensity and safe exercise progression (**45). **It was important to develop independent engagement with exercise during the program.** Toward the end of the program, care coordinators explored local provision to support transition to local fitness facilities for continuation of participants' exercise regimens at program termination |

#### CMOC 9 ('Using tools for self-management')

**When individuals with SMI and weight gain from anti-psychotic medications employ data from easy-to-use, convenient, lifestyle tracking tools and self-monitoring devices (e.g., fitbits) to monitor the impacts of their behaviours (C), individuals are more motivated to attain and maintain lifestyle changes (O), due to increased self-awareness of how certain behaviours (e.g., diet, exercise) can positively or negatively influence their health (M).**

CMOC based on data extracted from 20 documents.

| Document (author, date) | Extracted data |
|---|---|
| (Aschbrenner et al., 2016) | The wearable physical activity trackers (Fitbits) were a motivator for participants and their [exercise] partners to increase their physical activity, even among those who were not able to work out together. **Overall, participants reported high satisfaction with using the activity tracking devices, stating the devices were easy to use, motivational, and helpful for setting and monitoring goals**. In many cases, participants reported a friendly competition with their partner for increasing daily steps. One participant commented, "We were competing against each other because we had the Fitbit. It was fun. I would do it again." |
| (Casagrande et al., 2010) | **Self-monitoring and positive reinforcement** are important aspects of successful weight loss trials. Participants are asked to fill out a "Tracker" as a self-monitoring tool outside of group sessions. Each tracker is used for one week; participants record the number of servings of fruits and vegetables, and respond yes or no to: exercising for 30 minutes; drinking sugar drinks; eating junk food; smart portions or smart snacks. **The Tracker provides a behavioural cue to participants**. |

| | |
|---|---|
| (Stefancic et al., 2021) | For most participants, the trial-and-error process of self-monitoring began with checking the calorie content of familiar foods, resulting in feelings of shock. **This led many to develop a greater understanding** of the role of food choices in weight gain and the need to change food habits. **Self-monitoring of physical activity often increased awareness of current activity levels and the need to walk more.** Despite initial interest, few participants continued self-monitoring, citing challenges including that it was time consuming, not useful, or "too complicated." The findings suggest that future interventions should expand support for self-monitoring, meal planning, tailored physical activity, and advocacy. Such interventions should also enhance **participants' understanding of the corresponding impact of changes on weight loss and emphasize subjective wellness outcomes to maintain motivation** |

#### CMOC 10 ('Succeeding with family and friend support')

**When individuals with SMI and weight gain from anti-psychotics have family members/friends who emotionally support them with encouragement, praise and recognition; provide practical support (e.g., transportation); and/or actively participate with them in weight management activities (C), individuals are more apt to achieve and sustain their behaviour change goals (O), due to increased social reinforcement for their lifestyle change efforts (M).**

CMOC based on data extracted from 13 documents.

| Document (author, date) | Extracted data |
|---|---|
| (Aschbrenner et al., 2013) | **Participants cited empathy, validation, praise and encouragement from family members and significant others as supporting their efforts to achieve their health goals**. *One participant described receiving praise from friends when she lost weight: …"to the gym but she is going so she'll say, 'Why don't you come along,' and I eventually do go, and I feel better afterwards. And I've said, 'Why don't you come with me?,' just so she'll go more consistently. So we've helped each other.''* |
| (Aschbrenner et al., 2016) | **Participants viewed emotional support (e.g., encouragement, praise, recognition) and practical help (e.g., transportation) for health behaviour change as potential benefits of involving family members** and friends in a lifestyle intervention along with the potential to enhance the quality of these relationships (Aschbrenner et al., 2012)…One support partner commented**, "I think it brought us together—when you are with somebody and you are exercising, you can talk more and get whatever is off our chest, so I think it worked."** |
| (Pedley et al., 2018) | . *…my mum's been very, like, ringing me up going, are you still eating healthy, and stuff, and that's* ***actually got me close to my mum again*** *because me and my mum have been very distant…"* (Pt.2)<br>***…"they [friends] sort of changed with me, to sort of show well, we can do it as well, so you can."*** (Pt.14) |

#### CMOC 11 ('Facilitated, peer group participation')

**When individuals with SMI and weight gain from antipsychotic medications participate in facilitated, community-based, small-group weight management programs with similar group participants in a psychologically safe, non-stigmatizing space (C), individuals are more likely to engage in activities and learn important social skills, such as co-operation and accountability (O), due to a sense of belonging among peers (M).**

CMOC based on data extracted from 36 documents.

| Document (author, date) | Extracted data |
|---|---|
| (Gossage-Worrall et al., 2019) | There was an overwhelming sense that service users found the peer group interaction and social support to be beneficial. Some service users had never met others with schizophrenia and had attempted weight loss at commercial programmes but did not want to talk about the impact of their medication because of stigma attached to severe mental illness. **"I find it fantastic … meeting other groups of people on a similar medication with similar problems**..." (Service User S01/Q01) |
| (Green et al., 2015) | *"I really enjoyed the group setting. And I could sit next to anybody in the class and be perfectly comfortable. Because we all shared this kind of common mental health issue … I really liked the support …. You know, how did you do this week? What were your successes? What were your failures? "The part I didn't like was when the group setting ended. That was hard for me. I tried to go out and find another group … like Weight Watchers, and I couldn't find a group that I clicked with. So it was really frustrating to have that camaraderie and then lose it."* |
| (Nover, 2013) | All of the participants who engaged with the CalMEND CPCI program reported that they found the activities involving groups to be helpful because of the support the group provided…. **Participants reported that the group atmosphere felt inclusive and nonjudgmental, which helped them make progress toward their goals**… Elizabeth and Anna respectively describe this phenomenon: *Everybody I've met is sort of like me. They all have the same problems ... so that we all share ...* ***we can all share and no one's critical, so that makes it safe.*** *Because when I leave [the CalMEND program], I just feel my self-esteem is just on cloud nine. I just feel comfortable. ... And not feel like you're gonna be judged.*<br><br>Samantha, who earlier reported feeling like she has been treated like a number in her medical care, describes the group benefits of CalMEND: *"... it's been helpful for me about the foods and then self esteem, that self esteem class. And just expressing your feelings. And just seeing how everybody else is doing too, and getting their input. That's important to have a social network of a support group system."..*<br>[Anna]: *"Everybody I've met is sort of like me. They all have the same problems ... so that we all share ... we can all share and no one's critical, so that makes it safe. Because when I leave [the CalMEND program], I just feel my self-esteem is just on cloud nine. I just feel comfortable. ... And not feel like you're gonna be judged."* |
| (Park et al., 2017) | *"We keep each other on track, asking each other what exercise we have done that week…**I felt more determined to stay on track** then" (Rose) and "I really enjoyed walking together each week" (Clover).* |

| | |
|---|---|
| | *One participant noted increased exercise changes that she made after attending the group each week: "So now I go on another walk as well, two walks each week" (Hyacinth) and "when the weather changes I use an exercise DVD, so I still get my exercise each week" (Hyacinth).* |
| (Roberts & Jois, 2013) | Participants' need to establish friendships, bond with staff, feel a **sense of belonging, and gain support and encouragement from other**s was a key issue. Being part of a group, making friends, and **peer support from people in a similar situation** were central. […] Attending weekly sessions was viewed positively as a means of interacting socially with others. Littrell et al. (2003) attributed some of the benefits of their group-based intervention to the social support available. **Social networking** seems to be a key incentive for engaging people with SMI with group-based lifestyle interventions demonstrating that group activities are a useful format for delivering health promotion to this population: *I've tried weight-watchers and found that I didn't really get much out of going … you never make any personal relationships with people. … you* ***find that you can connect more with people here… having friendships here helps… I look forward to seeing people every week and that's what gives me that motivation to carry on coming****… (Participant 03*) |
| (Yarborough et al., 2016) | Several people noted that **accountability to self, to other group members, and to group leaders is what led to behaviour changes**. For example, one participant reported: "*Well, just knowing that I want to be accountable, because I don't want to disappoint the group or…myself, I guess"* (intervention arm, 3 months) |

#### CMOC 12 ('Trained peer mentor')

**When individuals with SMI and weight gain from anti-psychotic medications receive positive, unconditional support from appropriately trained, empathic, caring peers who openly share their own experiences and practical life skills (C), individuals are more likely to engage in and adopt healthier lifestyle behaviours (O) because peer supports' shared experiences are more credible to them (M) and there is an increased sense of hope (M) for more positive lifestyle outcomes.**

CMOC based on data extracted from 14 documents.

| Document (author, date) | Extracted data |
|---|---|
| (Bochicchio et al., 2021) | Peer providers often attended directly to participants' **sense of self-efficacy and were described as offering unwavering support and belief in participants' capacity for change.** These findings are in line with extant qualitative literature examining peer-led interventions that emphasize the **importance of fostering self-determination, and hope, all grounded in shared experience as a crucial means of effecting change in individuals with SMI** (Lietz et al., 2014*)…*<br>Participants viewed their relationships with peer and non-peer providers positively, but described differences in the approach to practice, power dynamics present, and how they identified with each provider. Participants described peers as process-oriented while |

| | |
|---|---|
| | non-peer staff as task-oriented, focusing on accomplishing concrete objectives. Each provider sought to boost participants' motivation, **but peers built hope by emphasizing the possibility of change, while non-peer providers emphasized the consequences of inaction**. Participants **related to peer staff through shared experiences, while identifying the importance of having a shared treatment goal with their non-peer provider.** Overall, participants appreciated the unique roles of both peer and non-peer staff in supporting their health… In addition to the ability of PS [peer support] staff to deliver intervention content, there was frequent emphasis on their ability to **cultivate a positive social environment, in which people felt comfortable sharing their own experiences and participating in the intervention:** *"He… makes a situation where…It's less intense… where you'd like to be and where you'd like to go.* ***And he makes you feel as though that you will achieve that*** *- just give yourself some time and relax and have fun while you're doing it…"* |
| (Bochicchio et al., 2019) | **"Caring"** was used to indicate that not only was the peer someone who was **truly concerned with their welfare**, but someone who also **understood the unique experience** of the participants and whose **efforts exceeded expectations**. Participants often described the peer-specialists as employees who came to work for *"more than just a paycheck."* One participant shared, ***"[The PS] cares... [The PS] goes out [their] way to make sure you alright…[the PS] ain't got to do that."*** *(IP7)* Another participant reiterated this sentiment, *"[The PS] really cares about how you're feeling. [The PS] knows what I'm going through. I hear voices sometimes, and [the PS] tells me that [they are] bipolar also. So, [the PS] got a similar...diagnosis."* (IP11) |
| (O'Hara et al., 2017) | Peers can use their own experiences to motivate clients to engage in healthy lifestyle changes and **serve as credible role models to support health behaviour change [11]…** Peer specialists' lived experience and **local knowledge were vital to develop practical adaptations and advice to support participants.** Peers' familiarity with participants' neighborhoods and socioeconomic limitations allowed them to share information about the best places to shop for healthy food as well as safe and affordable places to exercise. Their shared cultural backgrounds also enabled peers to provide practical advice on how to use healthier ingredients to prepare traditional dishes. Our findings suggest that peer specialists can **enhance the ecological validity of healthy lifestyle interventions for people with SMI** |
